# The avoidable health burden and healthcare costs related to alcohol consumption in Australia: multistate life table modelling

**DOI:** 10.1101/2024.12.12.24318952

**Authors:** Mary Njeri Wanjau, Linda Cobiac, Mishel Shahid, Amila Malawige, Leopold Aminde, Moosa Al Subhi, Phuong Nguyen, Mary Rose Angeles, Jaithri Ananthapavan, Lennert Veerman

## Abstract

**Background and Aims:** Excessive use of alcohol is one of the leading risks for mortality and disability globally. In Australia, alcohol was the fifth-highest risk factor contributing to disease burden in 2019. Estimates of the avoidable (future) alcohol related burden can help make the case for investment in preventive measures. This analysis aims to estimate the avoidable burden related to alcohol consumption in Australia.

**Design, setting, participants and intervention:** *The Alcohol Policy model (TAP),* a proportional multi-state lifetable model was developed and used to estimate the avoidable alcohol-related disease, injury and healthcare cost burden by comparing a scenario where the Australian adult population (aged ≥ 15 years) continues to drink alcohol at current rates to an identical population that consumes no alcohol. Taking 2020 as the base year, an open cohort was modelled over a 60-year time horizon.

**Measurements:** Changes in population alcohol consumption are modelled to lead to changes in 1) incident cases and mortality from alcohol-related diseases and injuries, 2) long-term health outcomes summarised as health adjusted life years (HALYs) and 3) healthcare costs. Results are reported in single years, over 25 years and 60 years (for HALYs and healthcare costs). No discounting was applied.

**Findings:** Over the first 25 years, elimination of alcohol consumption at the population level could prevent over 25.9 million incident cases of alcohol-related diseases and injuries (89% acute causes, 1% cancers and 10% other modelled chronic diseases). This translates to 5.1 (95% uncertainty interval [UI] 4.0 to 6.2) million HALYs gained and AUD 55 (95% UI 36 to 75) billion saved in healthcare costs. Over a 60-year period, the potential health benefits increase to 17 (95%UI 14 to 21) million HALYs and is associated with AUD 68 (95%UI 9.6 to 130) billion in healthcare cost-savings.

**Conclusion:** Our findings show that the avoidable alcohol-related disease, injury and healthcare cost burden in Australia is substantial. These findings reinforce the need for investment in effective and cost-effective polices that reduce alcohol consumption.

## INTRODUCTION

Excessive use of alcohol is one of the leading risks for mortality and disability globally accounting for approximately 2.4 million deaths and 92 million disability adjusted life years (DALYs) in 2019.^1,2^ In Australia, alcohol was the fifth-ranking risk factor in 2019, responsible for 5.1% of the total burden of disease and injury.^1^ In 2021, the social and economic costs of alcohol use were estimated to be $66.8 billion annually.^3^ According to the latest Organisation for Economic Co-operation and Development (OECD) report, Australians consume 9.5 litres/person/year.^4^The latest national estimates indicate that the alcohol-related disease burden is 1.9 times greater in the most disadvantaged socioeconomic group than in the least disadvantaged group.^5^ Consumption of alcohol impedes the achievement of UN sustainable development goals on equality, environment and economic and social development.^6,7^ Globally, alcohol use is the leading risk factor for mortality and disability for young adults aged 15-39 years.^1^ This age group bears most of the acute consequences due to high rates of injuries leading to death and disability.^8,9^ For people aged 40 years and above without underlying health conditions, consuming small amounts of alcohol (1 to 2 standard drinks per day) has been associated with possible protective effects for cardiovascular disease, stroke and type 2 diabetes mellitus.^8^ However, the evidence suggests there is no safe level of alcohol use for overall health.^2,3,8,10–12^ The risk of cancers and all-cause mortality rises with increasing levels of alcohol use.^8,10–13^ Hence, even small reductions in population-wide alcohol consumption levels can lead to a significant reduction in the health burden for individuals and the health system, and reduced costs for individuals, communities and governments.^3^ Reducing alcohol related harm is a global public health priority.^2,5,13–17^

Previous studies have looked at past exposures to alcohol consumption and estimated the attributable health and/or economic impact in Australia.^2,5,13–17^ Such estimates of attributable burden arising from past exposure are helpful in estimating future healthcare needs. Our study extends this previous work by illustrating how much of the future burden can be prevented (avoided) if action is taken now.

In this study, *The Alcohol Policy model (TAP)* was developed and used to assess the avoidable alcohol-related disease, injury and economic burden in Australia, i.e., the potential health benefits and cost savings that could be achieved if alcohol consumption was reduced at the population level.

## METHODS

### Overview

*The Alcohol Policy model* is developed in the R programming language (version 4.1.1). It uses proportional multi-state lifetable modelling methodology,^18^ an approach that has been widely used to evaluate population health and economic impacts of policy scenarios aimed at preventing chronic disease by addressing risk factors.^19–23^ To estimate the avoidable health and healthcare cost burden related to alcohol consumption, a modelled Australian population that continues to consume alcohol at current rates (business as usual) is compared to an identical population that has a lower level of alcohol consumption (counterfactual). In the main analysis, we model a ‘no alcohol use’ scenario to estimate the ‘avoidable burden’.^24^ We use *The Alcohol Policy model* to translate the changes in population alcohol consumption into changes in population health (in health adjusted life years [HALYs]), and future healthcare costs (in 2020 AUD) (Figure 1). In this section, each step of the modelling process is explained. In our reporting of both health and economic assessment aspects of our modelling we adhered to the Consolidated Health Economic Evaluation Reporting Standards 2022 (CHEERS 2022) statement: updated reporting guidance for health economic evaluations.^25^

**Figure 1:**
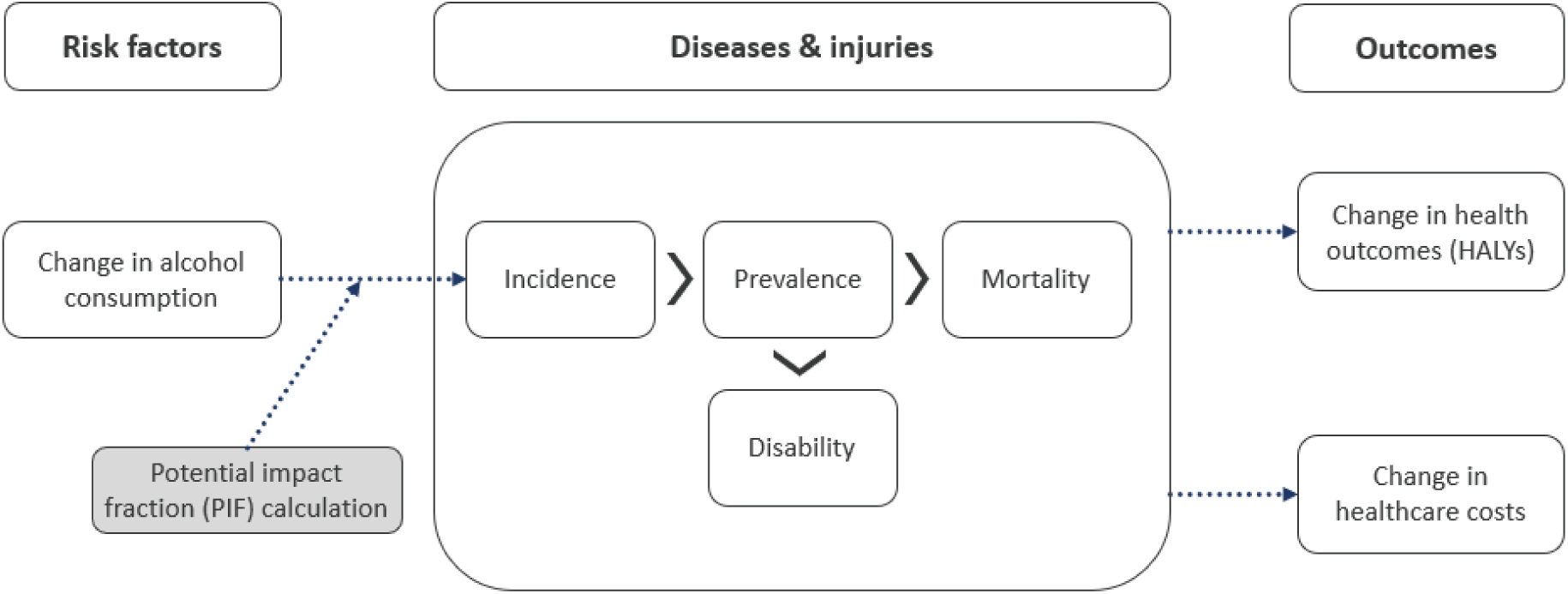
The analytical framework of quantifying the potential health benefits and cost impacts of changing alcohol use at the population level.

### Alcohol consumption data

In health surveys, alcohol consumption is typically underreported.^26,27^ To avoid underestimating alcohol consumption, we estimated a total envelope of volume (in litres) of alcohol purchased in Australia from Euromonitor 2019 sales data.^28^ Secondly, we adjusted the total volume consumed for an assumed 5% of waste (i.e., alcohol bought but not consumed).^29^ We multiplied this by the percentage of pure alcohol in each drink type (beer, cider, wine, spirits, ready-to-drink products and cocktails) in order to calculate the grams of pure alcohol consumed. Using population numbers^30^ and self-reported age and sex specific alcohol consumption recorded in the most recent National Drug Strategy Household Survey 2019 (NDSHS),^31^ we distributed the total estimated alcohol consumption across the population, by age and sex (Table 1). We applied a gamma distribution to the population alcohol consumption data to define the shape of the age/sex distribution of consumption.^26,32^

**Table 1:** Calculated alcohol consumption by age and sex for the year 2020.

| Age (years) | Calculated* quantity (grams/day) of pure alcohol consumed among drinkers |  |  |  | Prevalence of non-drinkers |  |
| --- | --- | --- | --- | --- | --- | --- |
|  | Female |  | Male |  | Female | Male |
|  | Mean | SD | Mean | SD | Percentage |  |
| 15-17 | 11.82 | 11.07 | 24.10 | 28.72 | 73.69% | 73.69% |
| 18-19 | 29.61 | 25.88 | 34.92 | 30.30 | 12.16% | 15.81% |
| 20-24 | 27.19 | 25.07 | 39.83 | 31.01 | 15.82% | 13.68% |
| 25-34 | 29.35 | 25.88 | 45.00 | 34.40 | 24.93% | 10.70% |
| 35-44 | 33.24 | 29.22 | 45.30 | 34.22 | 26.30% | 14.23% |
| 45-54 | 32.38 | 28.11 | 47.19 | 34.73 | 20.90% | 12.36% |
| 55-64 | 31.65 | 27.09 | 47.90 | 34.43 | 23.78% | 14.24% |
| 65-74 | 31.32 | 27.72 | 48.44 | 34.47 | 28.28% | 15.94% |
| 75-84 | 28.77 | 26.25 | 41.16 | 31.83 | 38.28% | 22.72% |
| 85+ | 28.77 | 26.25 | 41.16 | 31.83 | 41.77% | 30.07% |

\*See methods section for details on the calculation process and the data sources used

### The Alcohol Policy model

#### Development of the model structure and input data sources

The model uses male and female population cohorts in five-year age groups and simulates annual changes in health and mortality for each cohort until all have died or until a predefined time frame has elapsed (in this analysis, 60 years). To populate the model we used 2020 age and sex specific all-cause mortality rates from the Australian Bureau of Statistics^33^ and disability that is caused by all causes not explicitly included in the model (i.e., causes not associated with alcohol consumption) from the Global Burden of Disease (GBD) 2019^1,2^ (Supplementary File [SF] Table S1). Population numbers and projections for Australia were taken from the Australian Bureau of Statistics.^30,34^ We projected forward over a 60-year period based on the timeline for the State/Territory-specific lifetables projections.^34^ The model allows either an open or closed cohort analysis. In an open cohort, there is entry of new subjects based on the projected annual population growth rate for Australia.^33^ We carried out an open cohort analysis for this study.

We modelled all alcohol-related diseases and injuries based on dose-response relationships between alcohol consumption and disease or injury from meta-analyses undertaken for the GBD 2019 study.^35,36^ These included chronic diseases (ischaemic heart disease, ischaemic stroke, diabetes mellitus type 2, epilepsy, hypertensive heart disease, intracerebral haemorrhage, atrial fibrillation and flutter, larynx and pharynx cancer, lip and oral cavity cancer, liver cancer, breast cancer, colon and rectum cancer, oesophageal cancer, pancreatitis and cirrhosis)^35,36^ and acute causes (alcohol use disorders, interpersonal violence, lower respiratory infections, self-harm, transport injuries and unintentional injuries).^35,36^ For chronic alcohol-related diseases, we derived incidence, prevalence and case fatality from Global Burden of Disease data^1^ using DisMod II software (Figure S1)^37^ to estimate the epidemiological inputs to the model (Table S2).For acute alcohol-related causes, incidence, years lived with disability (YLD) and mortality rates were from the Global Burden of Disease data (Tables S3).^1^ In the lifetable analysis section below, we describe the modelling of the interaction between alcohol consumption levels and health outcomes.

The baseline healthcare costs for alcohol-related diseases and injuries, and the total per person costs of healthcare in Australia, were from the Australian Institute of Health and Welfare health expenditure data (an average of the 2018/19 and the 2019/20 reported costs).^38,39^ We derived the cause-specific unit costs by dividing the total cause-specific expenditure by the total number of disease/injury (cause) cases in a year as per the GBD data (Table S4). We calculated the average annual cost of all other healthcare (diseases not associated with alcohol consumption) by dividing the total remaining cost by the reported number of people in the Australian population (Table S5). The model takes account of changes in both the costs of alcohol-related diseases and injuries, and the costs of all other healthcare due to changes in life expectancy, i.e., costs incurred in added years of life caused by reduced alcohol consumption.^40,41^

#### Lifetable analysis

In the main analysis, we estimate the avoidable burden related to alcohol consumption by comparing a scenario where the Australian population continues to drink alcohol at current rates (with no trends applied) to a counterfactual population that consumes no alcohol.

Since the model is designed for evaluation of interventions aimed at preventing disease, it simulates the effect of an intervention (no alcohol consumption in the main analysis) on incidence (i.e., new cases) of disease. The change in incidence is calculated using the potential impact fraction (PIF), a measure of effect that calculates the proportional change in average disease incidence after a change in the exposure of a related risk factor.^42^ In this study, we apply the ‘distribution shift’ method in the PIF calculation (Equation 1).^42^

This method assumes a continuous alcohol consumption (risk factor) distribution (gamma distribution) with continuous relative risks (RRs) from GBD 2019^36^ modelled as lognormally distributed.^42,43^ We used the relative risk measures reported in GBD to simulate both the beneficial and harmful effects of alcohol consumption on diseases and injuries (Table S6).^35,36^ A relative risk below 1 indicates a beneficial effect and one above 1, a harmful effect. Although overall, the evidence suggests that there is no safe level of alcohol use for overall health,^2,3,8,10–12^ in the main analysis, we modelled beneficial effects to accommodate literature which shows that low and moderate levels of alcohol consumption confer a protective effect via ischaemic heart disease, ischaemic stroke, diabetes mellitus type 2.^13,35,36^ In the sensitivity analysis we included a scenario where the protective effects are excluded. For alcohol use disorders, which are entirely attributable to alcohol use, we assume the prevalence in drinkers goes up and down in proportion to quantity consumed.

The following PIF equation was applied:

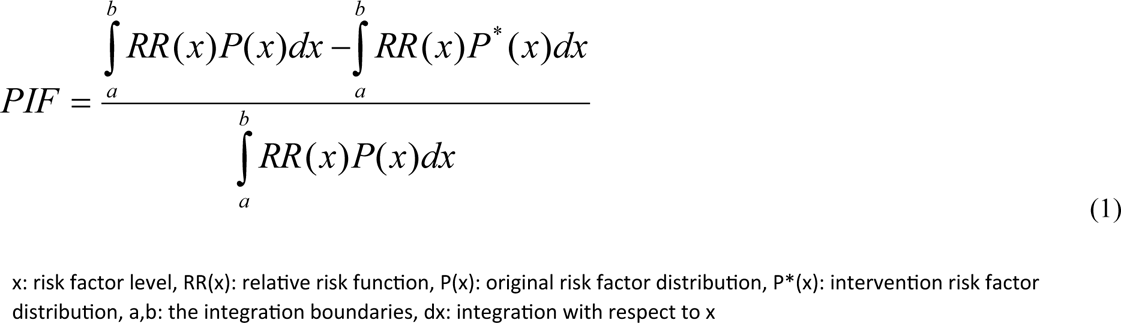

The change in disease incidence has flow-on effects on disease prevalence as the simulation time progresses; mortality, in turn, is modelled as a function of prevalence. Probabilities of dying and the size of the health adjustment (disability) are influenced by changing rates of incidence, prevalence and mortality of the diseases explicitly simulated in the model. Most diabetes-related deaths are from ischemic heart disease and ischemic stroke. To avoid double counting, we ignore diabetes mellitus type 2 mortality in the model. For the injuries, a change in the risk factor exposure (alcohol use) impacts the years lived with disability (non-fatal injuries) and mortality (Figure S2). Population health is measured in health-adjusted life years (HALYs) by adding up the years of life that are lived by the population, taking into account the years of healthy life lost due to disability per disease case (prevalent case for chronic and incident case for acute outcomes). Figure S2 shows a schematic description of how the disease-specific data are linked to the life table sections in a proportional multistate life table. Based on changing rates of diseases and injuries, the model simulates impacts on healthcare costs. The costs of healthcare for alcohol-related diseases and injuries included money spent by all levels of governments as well as non-government e.g., private health insurers, individuals and injury compensation insurers.^38,39^ We used areas of expenditure as defined in the Australian Institute of Health and Welfare health expenditure data, i.e., private hospital services, public hospital admitted patients, public hospital emergency department, public hospital outpatients, general practitioner services, specialist services, medical imaging, pathology, allied health and other services, and the pharmaceutical benefits scheme (prescriptions dispensed).^38,39^ We did not apply discounting in this analysis.

In Table 2, we provide a summary of the model input data, respective sources and where applicable, the distributions applied to account for uncertainty.

**Table 2:** A summary of model input data.

| Data | Source |
| --- | --- |
| Pure alcohol intake by age and sex | Euromonitor International<sup>28</sup> and 2019 National Drug Strategy Household Survey (NDSHS) (Table 1).<sup>31</sup> Gamma distribution applied. |
| Population numbers by sex and 5-year age group | National population data (2020) and projections over a 60-year period published by Australian Bureau of Statistics (Table S2).<sup>30,34</sup> |
| All-cause mortality rates by age and sex | Australian and State/Territory-specific lifetables published by the Australian Bureau of Statistics, 2020 data. (Table S2).<sup>33</sup> |
| Years of healthy life lost due to disability (YLD) rates by age and sex | All-cause and cause-specific rates of disability (YLD) were derived from GBD 2019 data (Table S2).<sup>1,2</sup> |
| Disease and injury rates by age and sex | Global Burden of Disease 2019 data (Tables S4 & S5).<sup>1</sup> We used DisMod II software to derive incidence, prevalence and case fatality inputs for chronic alcohol-related diseases. |
| Healthcare costs by age and sex (disease specific costs & the average annual cost of *all other healthcare*) | Australian Institute of Health and Welfare health expenditure data (an average of the 2018/19 and the 2019/20 reported costs, Table S7).<sup>38,39</sup> Lognormal distribution applied. |
| Dose-response functions for each alcohol-related disease or injury | Relative risks of disease and injury due to alcohol intake from the Global Burden of Disease study 2019<sup>36</sup> (Table S3). Lognormal distribution applied. |

### Uncertainty and sensitivity analysis

We conducted uncertainty analysis using Monte Carlo simulation (bootstrapping) in which we modelled input variables with a known distribution. This included distributions in alcohol consumption at baseline, relative risks linking exposure to disease/injury incidence, and healthcare costs (Table 2). We ran the model 2,000 times, drawing new random values for each input variable for each run. The model output reflects the joint uncertainty associated with the parameters that were varied (‘parameter uncertainty’). We report the outputs as mean point estimates and 95% uncertainty intervals (UI) defined as the 2.5^th^-97.5^th^ percentiles, respectively, based on the 2000 runs of the Monte Carlo simulation. We conducted the following scenario analyses to test how important variables influenced the results: i) excluding the beneficial/protective effects of low and moderate alcohol consumption,^8^ and ii) population meeting the current guidelines, modelled as a maximum of 10 standard drinks in a week where one standard drink is 10 grams of pure alcohol).^9^

## RESULTS

We present results for the theoretically avoidable alcohol burden (if exposure could be suddenly changed to optimal levels, i.e., no alcohol consumption).

### Changes in incident cases of disease and injuries

Over the first 25 years (2020 to 2045), approximately 25.9 million incident cases of diseases and injuries (23.1 million acute causes, 276,857 cancers and 2.6 million other modelled chronic diseases) could be prevented if alcohol consumption was eliminated in the Australian population (Table 3). Of the acute causes included in the model, the greatest reduction in incident cases was seen in unintentional injuries (12.7 million, 95%UI 3.1 to 22.5) followed by alcohol use disorders (6.5 million) (Figure S3). Of the modelled cancers, the greatest reduction was estimated for colon and rectum cancer (111,523, 95%UI 75,673 to 146,459) followed by breast cancer (female) (81,497, 95%UI 47,395 to 117,433). Of the other modelled chronic diseases, the greatest reduction was predicted for cirrhosis (2.3 million, 95%UI 2.0 to 2.5) followed by atrial fibrillation and flutter (221,484, 95%UI (161,067 to 282,644). Overall, a greater avoidable burden was predicted in males than females. In this main analysis, the protective effects of low and moderate alcohol consumption on diabetes mellitus type 2, ischemic heart disease and Ischemic stroke are included. The protective effect partly offsets the reduction in incident cases with results showing an overall slight increase of incidence of diabetes mellitus type 2 and ischemic heart disease cases (Table 3). In addition, some results show negative values (additional incident cases). This can be explained by first, the putative protective effect of low-dose alcohol and second, disease and injury cases that arise in added years of life (people who live longer due to reduced alcohol mortality are still at risk of these conditions).

**Table 3:** Incident cases that could be prevented over the first 25 years (2020 to 2045) if alcohol consumption was eliminated in the Australian population.

| Alcohol related diseases & injuries | Mean, 95%UIs |  |  |
| --- | --- | --- | --- |
|  | Female | Male | Total |
| **Acute causes** |  |  |  |
| Alcohol use disorders | 2,195,385 | 4,335,403 | 6,530,788 |
|  | (2,195,385 to 2,195,385) | (4,335,403 to 4,335,403) | (6,530,788 to 6,530,788) |
| Interpersonal violence | 181,494 | 702,815 | 884,310 |
|  | (41,739 to 312,183) | (189,886 to 1,148,286) | (366,021 to 1,360,188) |
| Lower respiratory infections | 611,682 | 1,527,079 | 2,138,761 |
|  | (-93,251 to 1,297,791) | (401,829 to 2,641,884) | (773,166 to 3,433,016) |
| Self-harm | 63,944 | 83,445 | 147,389 |
|  | (11,596 to 111,716) | (32,466 to 128,083) | (77,816 to 216,139) |
| Transport injuries | 234,217 | 418,239 | 652,457 |
|  | (48,780 to 412,197) | (111,732 to 687,131) | (305,799 to 978,160) |
| Unintentional injuries | 4,156,918 | 8,575,466 | 12,732,384 |
|  | (54,479 to 8,317,488) | (-292,410 to 17,051,559) | (3,118,138 to 22,455,727) |
| **Cancers** |  |  |  |
| Breast cancer (female) | 81,497 |  | 81,497 |
|  | (47,395 to 117,433) |  | (47,395 to 117,433) |
| Colon and rectum cancer | 36,763 | 74,760 | 111,523 |
|  | (19,457 to 53,544) | (42,883 to 105,311) | (75,673 to 146,459) |
| Larynx and pharynx cancer | 3,956 | 20,518 | 24,474 |
|  | (2,535 to 5,279) | (15,418 to 24,926) | (19,210 to 29,077) |
| Lip and oral cavity cancer | 6,493 | 20,232 | 26,725 |
|  | (4,873 to 7,889) | (17,067 to 23,128) | (23,138 to 29,979) |
| Liver cancer | 1,918 | 5,949 | 7,867 |
|  | (-402 to 4,221) | (-389 to 11,814) | (1,182 to 14,030) |
| Oesophageal cancer | 5,638 | 19,134 | 24,772 |
|  | (3,308 to 7,780) | (13,074 to 24,286) | (18,631 to 30,361) |
| **Other chronic diseases** |  |  |  |
| Atrial fibrillation and flutter | 78,001 | 143,483 | 221,484 |
|  | (45,968 to 111,340) | (88,706 to 195,546) | (161,067 to 282,644) |
| Cirrhosis | 862,359 | 1,416,718 | 2,279,077 |
|  | (630,449 to 1,031,881) | (1,181,987 to 1,587,654) | (1,975,107 to 2,532,381) |
| Diabetes mellitus type 2 | -96,470 | 59,579 | -36,891 |
|  | (-266,530 to 78,705) | (-115,782 to 225,786) | (-279,513 to 208,483) |
| Epilepsy | 24,307 | 38,623 | 62,931 |
|  | (13,506 to 33,874) | (24,402 to 51,227) | (45,407 to 79,700) |
| Hypertensive heart disease | 17,410 | 17,552 | 34,963 |
|  | (3,750 to 30,698) | (6,548 to 27,537) | (17,142 to 51,446) |
| Intracerebral haemorrhage | 10,579 | 17,677 | 28,256 |
|  | (1,919 to 19,079) | (10,058 to 25,045) | (16,793 to 38,832) |
| Ischaemic heart disease | -86,736 | -99,370 | -186,106 |
|  | (-242,660 to 70,846) | (-650,104 to 425,468) | (-744,135 to 350,779) |
| Ischaemic stroke | -7,816 | 24,497 | 16,680 |
|  | (-56,953 to 39,445) | (-12,518 to 59,309) | (-44,829 to 74,104) |
| Pancreatitis | 54,652 | 102,628 | 157,280 |
|  | (18,843 to 84,495) | (66,866 to 130,867) | (106,625 to 201,843) |

Main analysis scenario modelled: Alcohol consumption eliminated in the Australian population with protective effects of low and moderate alcohol consumption on ischemic heart disease, diabetes mellitus type 2 and Ischemic stroke included. Negative values can be explained by the questionable alcohol protective effect in the relative risk measures and second, disease & injury cases that arise in added years of life (people live longer due to reduced alcohol harm and they are still at risk of these diseases and injuries).

### Changes in mortality

Over the first 25 years, the model predicted that approximately 211,697 deaths could be averted if alcohol consumption was eliminated in the Australian population. About 82,093 of these deaths were from acute causes, 114,948 from cancers and 14,656 the other chronic diseases modelled. The greatest reduction from acute causes was related to self-harm followed by alcohol use disorders (Table 4 & Figure S4).

**Table 4:**
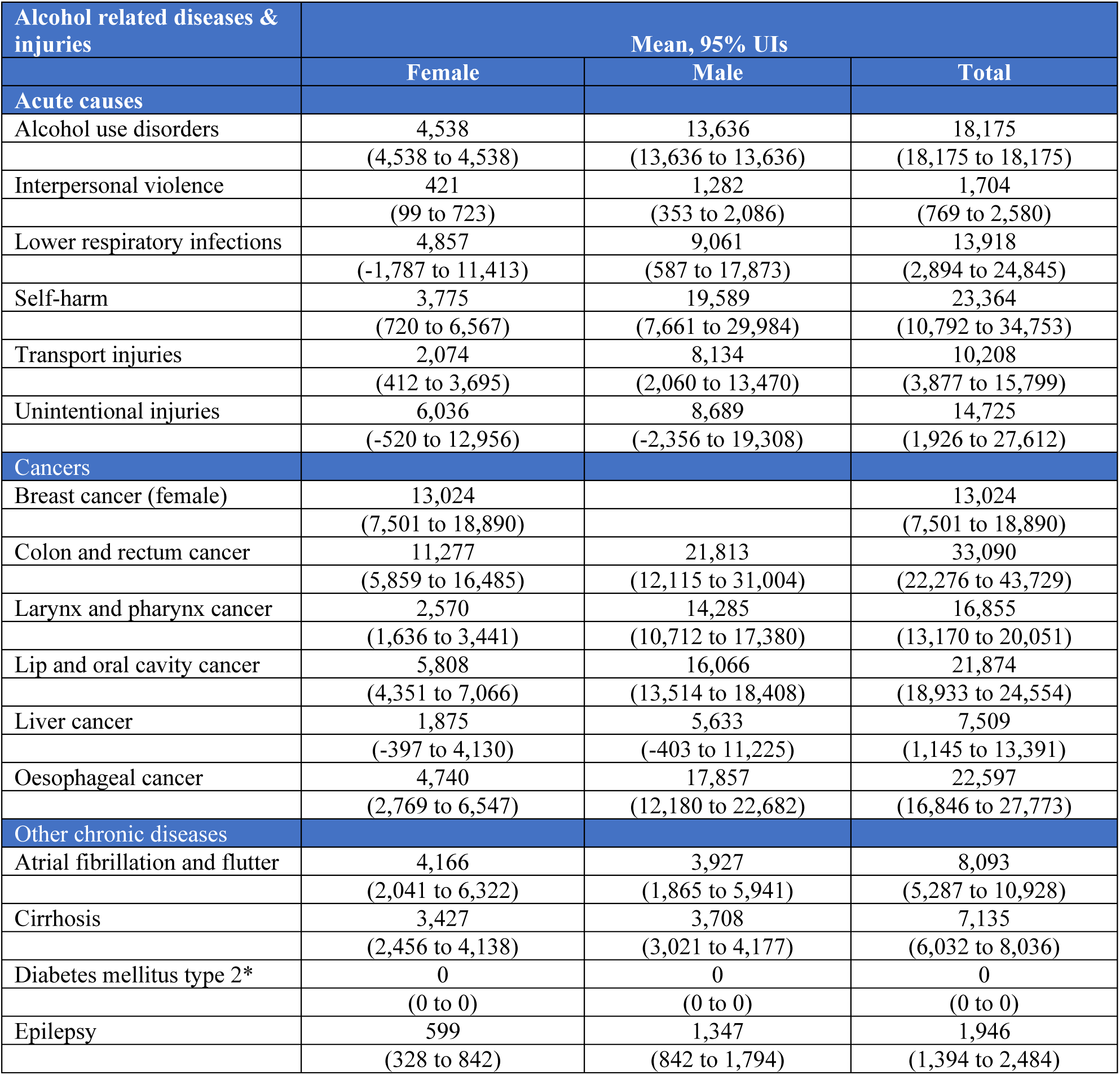
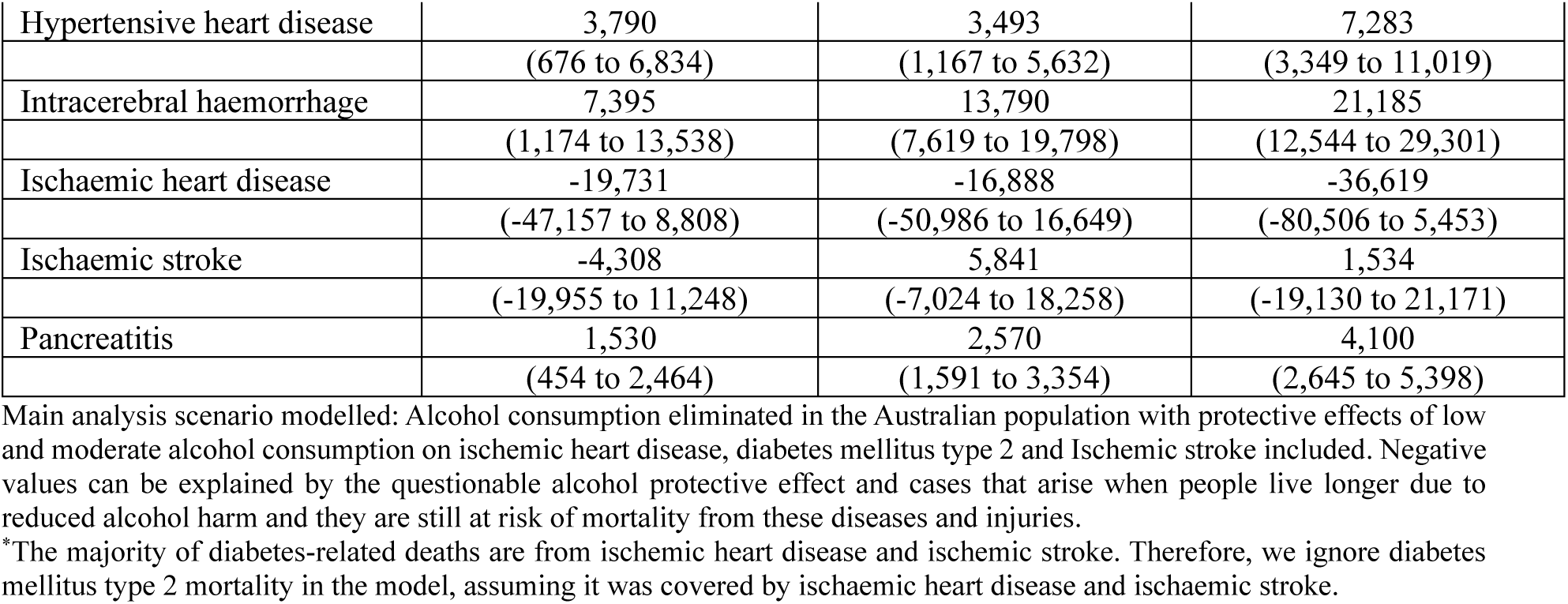
Cause-specific deaths that could be averted over the first 25 years (2020 to 2045) if alcohol consumption was eliminated in the Australian population.

As shown in Table 4, the mortality reductions modelled were partly offset by the protective effect modelled via ischemic heart disease and ischemic stroke. As with incidence, negative values can be explained by the putative alcohol protective effect and cases that arise in added years of life.

### Changes in health adjusted life years (HALYs)

Over 25 years’ time, an estimated 5.1 million (95%UI 4.0 to 6.2) HALYs could be saved. Over 60 years’ time (2020 to 2080), the model predicted that elimination of alcohol consumption at the population level could save approximately 17.0 million (95%UI 13.7 to 20.6) HALYs. Figure S5 shows the HALYs gained over time. Greater gains were estimated in males than females.

### Changes in healthcare costs by area of expenditure

When both the changes for costs of alcohol-related diseases and injuries and the costs of all other healthcare are modelled, reducing alcohol consumption to zero could save approximately AUD 55 billion (95%UI 36 to 75) in the first 25 years. Over 60 years, approximately AUD 68 billion (95%UI 9.6 to 130) could be saved. The results for over 60 years are greatly impacted by the inclusion of the costs of all other healthcare (i.e., costs incurred in added years of life since a reduction of alcohol consumption prolongs life). Figure 2 shows the estimated changes per the areas of expenditure defined by the Australian Institute of Health and Welfare. Chart A shows for when only the changes in the costs of alcohol-related diseases and injuries are modelled. Chart B shows changes for both the costs of alcohol-related diseases and injuries and the costs of all other healthcare.

**Figure 2:**
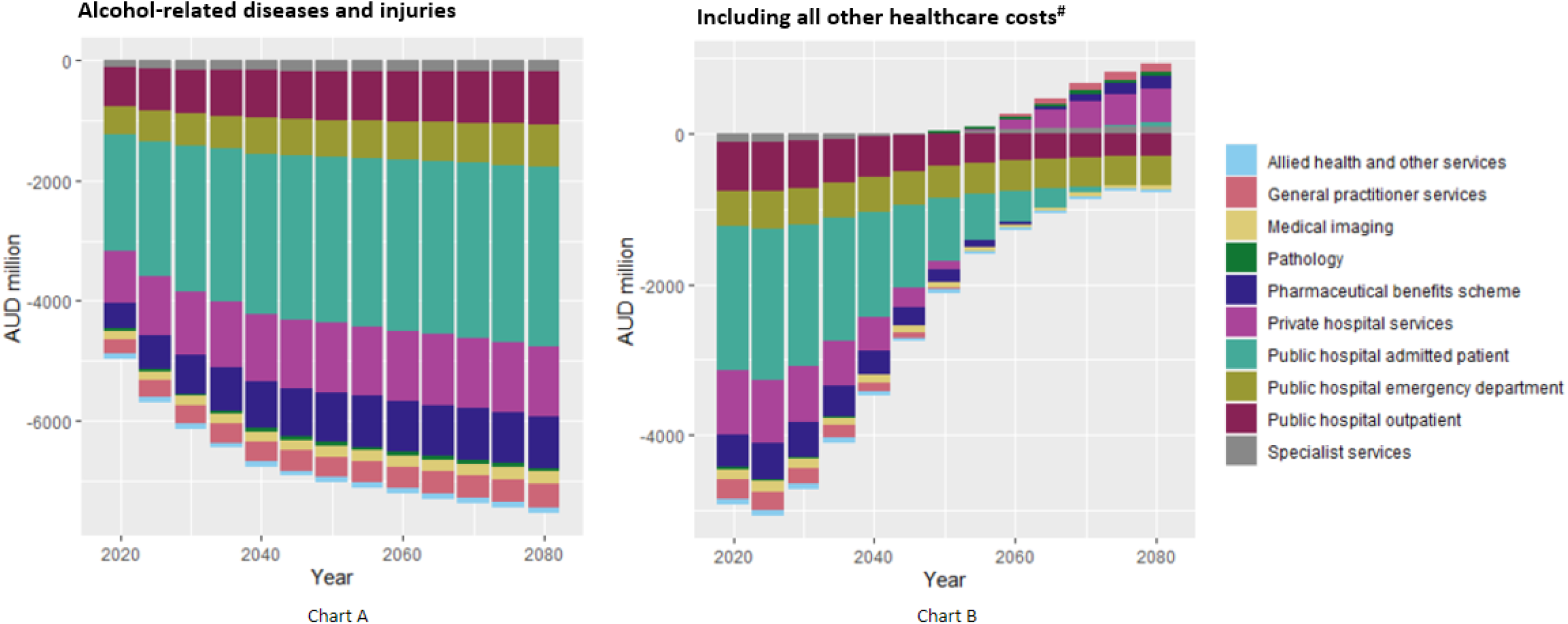
Healthcare cost changes over time if alcohol consumption was eliminated in the Australian population. The estimated changes are presented per the areas of expenditure defined by the Australian Institute of Health and Welfare.^38,39^ Chart A shows changes for when only the changes in the costs of alcohol-related diseases and injuries are modelled. ^#^Chart B shows changes for both the costs of alcohol-related diseases and injuries and the costs of all other healthcare due to changes in life expectancy, (i.e., costs incurred in added years of life since a reduction of alcohol consumption prolongs life).^40,41^

### Sensitivity analysis results

When compared to the main analysis, excluding the beneficial/protective effects of low and moderate alcohol consumption resulted in an increase in the preventable incident cases and deaths (Table S7). This translated to modest increases in HALYs gained in both 25- and 60-year time horizons (Table 5 & Table S8) and healthcare costs saved in 25-year time horizon (Table 5). Over 60 years’ time, the healthcare cost savings are reduced by the inclusion of costs incurred in added years of life.

**Table 5:** Changes in HALYs and healthcare costs for alternative scenarios modelled.

| Scenario/ Variable | Years | HALYs gained<br>(Million) | Healthcare cost savings *<br>(Million) |
| --- | --- | --- | --- |
|  |  | Mean (95% UI) | Mean (95% UI) |
| Avoidable burden (main analysis) | 00-25 | 5.1 (4 to 6.2) | 55,000 (36,000 to 75,000) |
|  | 00-60 | 17 (14 to 21) | 68,000 (9,600 to 130,000) |
| Avoidable burden with protective effects excluded* | 00-25 | 5.5 (4.6 to 6.5) | 56,000 (38,000 to 74,000) |
|  | 00-60 | 19 (16 to 22) | 60,000 (5,900 to 110,000) |
| Met alcohol consumption guidelines | 00-25 | 4.4 (3.2 to 5.5) | 49,000 (30,000 to 69,000) |
|  | 00-60 | 15 (12 to 19) | 56,000 (-1,700 to 110,000) |

\*Healthcare costs in 2020 AUD. Scenarios modelled: Avoidable burden: Population level elimination of alcohol consumption (main analysis); Avoidable burden with protective effects excluded: Population level elimination of alcohol consumption with protective effects excluded (assessing sensitivity to excluding the beneficial/protective effects of low and moderate alcohol consumption on ischemic heart disease, diabetes mellitus type 2 and Ischemic stroke)<sup>8</sup>; Met guidelines: Population met the current alcohol consumption guidelines (modelled as a maximum of 10 standard drinks in a week where one standard drink is 10 grams of pure alcohol)<sup>9</sup>; Healthcare costs are in AUD 2020.

Over the first 25-year time horizon, an estimated 4.4 (95%UI 3.2 to 5.5) million HALYs could be saved if the Australian population met the current alcohol consumption guidelines (Table 5 and Table S8). This is 14% less than the avoidable burden estimated for the main analysis where we model no alcohol consumption in Australia. In the same period, AUD 49 (95%UI 30 to 69) billion in healthcare costs could be saved, 11% less than the savings seen in the scenario where alcohol consumption is eliminated (Table 5). Figures S5 and S6, show changes in HALYs and healthcare costs over different time periods for various modelled scenarios.

## DISCUSSION

### Statement of principal findings

In this study, we found that elimination of alcohol consumption at the population level could lead to prevention of over 25 million incident cases of diseases and injuries over the first 25 years. This translated to 5.1 million HALYs gained and AUD 55 billion saved in healthcare costs. Over 60 years’ time, 17 million HALYs could be gained and AUD 68 billion could be saved. The inclusion of the costs of all other healthcare (i.e., costs incurred in added years of life) reduces the costs savings over time. Overall, a greater avoidable burden was seen in males than females. This is mainly due to higher baseline alcohol consumption levels.^31^ In addition, for most alcohol related conditions modelled, there was greater incidence and mortality in males compared to females.^1,2^

Epidemiological models of alcohol have been developed previously^44–47^ but to our knowledge, ours is the first to estimate the avoidable burden for Australia. A previous Australian study did report that if the annual adult alcohol consumption was reduced by 3.4 litres/capita, the annual numbers of new cases of alcohol-related disease, deaths and disability adjusted life years could be reduced by 98,000, 380 and 21,000, respectively, for the 2008 Australian population.^44^ Internationally, three previous studies have estimated the avoidable future burden of alcohol use for the Nordic countries and Canada.^45–47^ In the Nordic study, the authors found that over a 30-year period, if alcohol consumption was eliminated, 83,000 cancer cases could be avoided.^45^ Similar to our study, the number of avoidable cancer cases was highest in breast cancer and colon and rectum cancer. In a Danish study, authors found that if population alcohol consumption was immediately reduced to the recommended level (below 12 grams/day), 445 new cases of breast cancer could be prevented in 45 years’ time.^46^ The Canadian study included estimation of the future avoidable cancer burden under four counterfactual scenarios that included instantaneous lowering of alcohol consumption meeting the World Cancer Research Fund (WCRF) low risk guidelines (≤1 drink/day for females; ≤2 drinks/day for males) and meeting the Canada’s Low-Risk Drinking Guidelines (≤2 drink/day for females; ≤3 drinks/day for males).^47^ They found that over the first 25 years, 36,536 incident cancer cases could be avoided if the WCRF guidelines were met while 18,365 cancer cases could be avoided with compliance to the Canadian low-risk alcohol drinking guidelines.^47^ These numbers are not directly comparable to our study due to country differences in demographics, baseline alcohol consumption levels and disease rates. Methodological differences included the modelling of alcohol consumption in defined categories whereas our study used continuous distributions of consumption in grams of alcohol by age and sex. In addition, these previous studies apply various latency and/or lag times to allow for delayed changes in disease risk after changes in exposure to risk factor (alcohol consumption).^45–47^ In our study, we use the lifetable method which deals with lag time for chronic diseases by modelling the impact on incidence, which leads to changes in prevalence, which in turn affects mortality.^18^

### Strengths and limitations

A strength is that we used an established robust proportional multi-state lifetable approach and assessed both the avoidable health burden related to alcohol consumption in Australia, and health care cost savings.

In the model, we take into account expected future population numbers and all-cause mortality rates as projected by the Australian Bureau of Statistics.^33^ We also included scenario analyses to test how important variables influenced the results. We incorporate uncertainty for the level of alcohol consumption, relative risks and healthcare costs. However, the 95% uncertainty intervals in our results underestimate the true level of uncertainty. Uncertainty around the disease incidence and prevalence input data was not modelled for technical reasons. As detailed in methods section, we use DISMOD II to ensure internal consistency of the disease parameters, and DISMOD does not allow for uncertainty around estimates. Furthermore, we assumed that disease and injury rates would remain constant into the future and only applied the change resulting from the modelled counterfactual scenario. We also did not model trends in population’s alcohol consumption levels. Future studies could incorporate these trends or projections. In determining the current intake of pure alcohol by age and sex in Australia (baseline consumption levels in the model), we applied adjustments for underreporting often seen in health surveys,^26,27^ for alcohol bought but not consumed^29^ and for the percentage of pure alcohol by drink type. Due to lack of data, we did not adjust for unrecorded alcohol, i.e., alcohol produced, distributed and sold outside the formal channels under government control.^6,48^ Unrecorded alcohol is estimated to be approximately 9.4% for high income countries.^49^ Underestimation of the alcohol consumption level in our model could result to underestimation of the avoidable burden.

Moreover, alcohol consumption harm is experienced by not only the person drinking but often also by others, including those who abstain from drinking.^50,51^ Examples of such harm include drunk driving, foetal alcohol syndrome disorder, noise, fear of physical abuse, etc.^51^ The estimated avoidable burden in this study does not include alcohol harm caused to others. The relative risk estimates we used to model the association of consumption levels with health outcomes may suffer from residual confounding despite adjustments for confounding and bias impacts done in the GBD study.^8^

### Research implications on policy and practice and future research

Our findings can help policy makers to understand the scale of the future alcohol burden that can be avoided. The scenario where population meets the current alcohol consumption national guidelines is insightful for preventive action in Australia where, notwithstanding a downward trend, one in three adults consume alcohol in excess of the guidelines.^52^

Our findings confirm that there is no safe level of alcohol use for overall health and the health risks rise the more a person drinks.^2,3,8–12^ In our main analysis, the putative protective effect of low/moderate alcohol consumption via cardiovascular disease and diabetes mellitus type 2 for older ages^35,36^ is outweighed by an increased risk of injuries, cancers and all-cause mortality.^13^

Our findings underscore the consequences of inaction and reinforce the health and economic case for preventive measures, especially for Australia, where alcohol is the most widely used drug.^9^ Our analysis did not assess impacts by population subgroups, but it seems likely that alcohol control can reduce health inequities.^5,53^ The avoidable burden for people with low socio-economic status may be disproportionally high since they may be more vulnerable to alcohol related harm due to factors that could increase their risk of injury, such as nutrition deficiencies and unsafe drinking settings. Future research could estimate the avoidable alcohol related burden in Australia by social economic grouping and ethnicity, e.g., First Nations and culturally and linguistically diverse populations.

Australia’s National Alcohol Strategy 2019-2028 outlines the national commitment to reducing alcohol related harm through both early intervention and healthcare strategies, and through prevention and law enforcement strategies.^54^ Our findings support prioritisation of investment in alcohol harm reduction. Policies and interventions that reduce consumption at the population level are likely to offer favourable impact^55^ as they create environments that support and/or allow for an increase in the number of people who abstain from alcohol consumption, a delay in the age when people start drinking, and reductions in alcohol consumption for those who drink.

Evidence on effective and cost-effective population level policies and interventions that reduce alcohol consumption could inform the priority setting and selection of policies for implementation. In further work we will be using *The Alcohol Policy* model to estimate the long-term health, cost impacts and cost effectiveness of selected policies that reduce the consumption of alcohol in Australia.

### Conclusion

Our study shows that elimination of alcohol consumption in Australia could lead to prevention of over 25 million incident cases of diseases and injuries over the first 25 years translating to 5.1 million HALYs gain and AUD 55 billion savings in healthcare costs. Stronger alcohol control policies are needed to realise these gains.

## Acknowledgements

The development of *The Alcohol Policy model (TAP)* was part of a larger project at the national level and for the Western Australia State. We acknowledge the stakeholders and policy partners who were engaged in the research project. This was a project with The Australian Prevention Partnership Centre (TAPPC) and Western Australia Mental Health Commission. We acknowledge the Australian Institute of Health and Welfare (AIHW) as the source of the alcohol consumption data from the National Drug Strategy Household Survey 2019 (NDSHS).

## Author contribution

**Mary Njeri Wanjau:** Conceptualization (equal); Investigation (equal); Data curation (equal); Formal analysis (supporting); Writing – original draft preparation (lead); Visualization (equal); project administration (lead); Writing – review & editing (equal).

**Linda Cobiac:** Conceptualization (equal); Investigation (equal); Data curation (lead); Software (lead); methodology (equal); Formal analysis (lead); Visualization (lead); Writing – review & editing (equal).

**Mishel Shahid:** Investigation (equal); Data curation (equal); Writing – review & editing (equal).

**Amila Malawige:** Writing – review & editing (equal).

**Leopold Aminde:** Visualization (equal); Data curation (supporting); Writing – review & editing (equal).

**Moosa Al Subhi:** Writing – review & editing (equal).

**Phuong Nguyen:** Writing – review & editing (equal).

**Mary Rose Angeles:** Writing – review & editing (equal).

**Jaithri Ananthapavan:** Supervision (supporting); Funding acquisition (equal); Writing – review & editing (equal).

**Lennert Veerman:** Conceptualization (equal); Investigation (equal); Data curation (supporting); methodology (equal); Formal analysis (supporting); Funding acquisition (equal); Supervision (lead); Writing – review & editing (equal).

## Data availability

The data that supports the findings of this study are available in the manuscript and supplementary material of this article

