## Supplementary Material for "The avoidable health burden and healthcare costs related to alcohol consumption in Australia: multistate life table modelling"

### Supplementary File

### Methods

#### Population, mortality and disability rates for Australia

*Table S1: Population and mortality rates for Australia by age & sex*

| **Age** | **Population** | | **Mortality rate*** | | **Disability (YLD) rate#** | |
|---|---|---|---|---|---|---|
| | Female | Male | Female | Male | Female | Male |
| 2 | 745,435 | 789,571 | 0.00010 | 0.00015 | 0.02100 | 0.02392 |
| 7 | 787,135 | 832,349 | 0.00007 | 0.00008 | 0.04104 | 0.03917 |
| 12 | 778,240 | 824,582 | 0.00009 | 0.00009 | 0.06413 | 0.05643 |
| 17 | 720,171 | 766,260 | 0.00019 | 0.00036 | 0.09017 | 0.07568 |
| 22 | 827,034 | 877,255 | 0.00023 | 0.00056 | 0.11697 | 0.09528 |
| 27 | 937,176 | 947,862 | 0.00028 | 0.00063 | 0.13312 | 0.10792 |
| 32 | 962,651 | 946,250 | 0.00042 | 0.00083 | 0.14082 | 0.11515 |
| 37 | 926,037 | 914,471 | 0.00061 | 0.00114 | 0.14628 | 0.12060 |
| 42 | 819,684 | 801,478 | 0.00092 | 0.00162 | 0.15194 | 0.12617 |
| 47 | 851,473 | 831,387 | 0.00141 | 0.00230 | 0.15781 | 0.13329 |
| 52 | 796,781 | 771,543 | 0.00211 | 0.00343 | 0.16489 | 0.14391 |
| 57 | 799,113 | 767,984 | 0.00310 | 0.00517 | 0.17582 | 0.16023 |
| 62 | 741,684 | 697,841 | 0.00450 | 0.00759 | 0.19252 | 0.18325 |
| 67 | 652,820 | 609,962 | 0.00701 | 0.01138 | 0.21455 | 0.21151 |
| 72 | 570,355 | 540,698 | 0.01184 | 0.01847 | 0.24172 | 0.24274 |
| 77 | 399,519 | 368,497 | 0.02122 | 0.03175 | 0.27410 | 0.27492 |
| 82 | 286,797 | 239,938 | 0.04071 | 0.05823 | 0.31126 | 0.30604 |
| 87 | 184,232 | 130,996 | 0.08161 | 0.10785 | 0.35399 | 0.33838 |
| 92 | 100,243 | 56,505 | 0.15549 | 0.18247 | 0.40252 | 0.37302 |
| 97 | 34,029 | 13,210 | 0.26071 | 0.27289 | 0.45676 | 0.40980 |
| Totals | 12,920,609 | 12,728,639 | | | | |

* All-cause mortality rates # rates of disability (YLD) that is caused by all causes not explicitly included in the model

Data sources: 2020 age and sex specific all-cause mortality rates, population numbers and projections (projected forward over a 60-year period) for Australia taken from the Australian Bureau of Statistics (ABS) data and State/Territory-specific lifetables,[1,2] and rates of disability (YLD) from GBD 2019 data.[3,4]

#### The conceptual disease model

Figure S1 shows the schematic description of how the disease-specific data are linked to the life table sections in a proportional multistate life table. It also shows the interaction with the disease models and the risk factor.

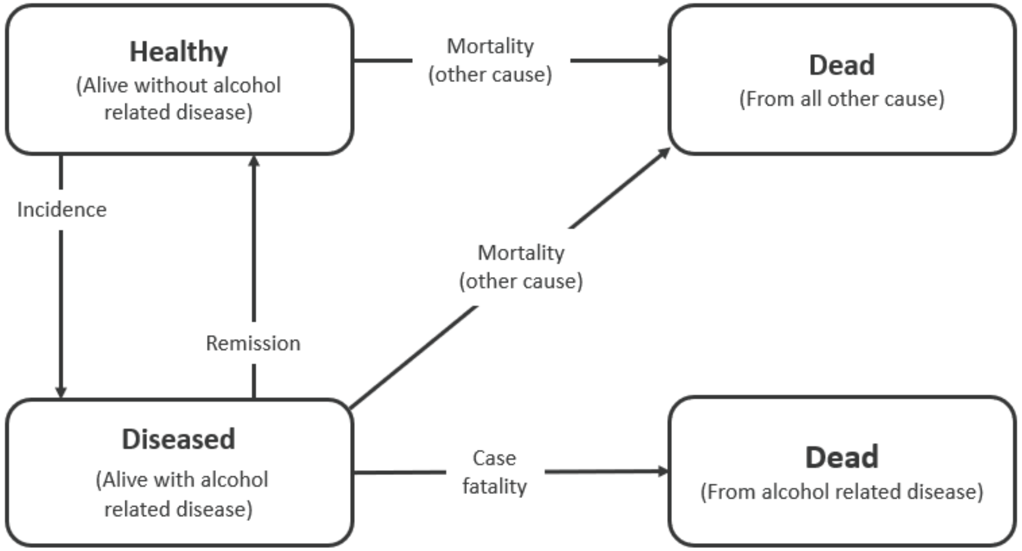

*Figure S1: The conceptual disease model showing four states and four transition hazards (adapted from Barendregt et al.[5])*

#### Disease inputs- Chronic diseases

*Table S2: Epidemiological data for chronic diseases modelled (rates per 1)*

| Cause | Age group (years) | Female | | | | | Male | | | | |
|---|---|---|---|---|---|---|---|---|---|---|---|
| | | Case fatality* | Incidence | Prevalence | Remission | YLD (disability) | Case fatality* | Incidence | Prevalence | Remission | YLD (disability) |
| Atrial fibrillation and flutter | 0-4 | - | - | - | - | - | - | - | - | - | - |
| Atrial fibrillation and flutter | 5-9 | - | - | - | - | - | - | - | - | - | - |
| Atrial fibrillation and flutter | 10-14 | - | - | - | - | - | - | - | - | - | - |
| Atrial fibrillation and flutter | 15-19 | - | - | - | - | - | - | - | - | - | - |
| Atrial fibrillation and flutter | 20-24 | - | - | - | - | - | - | - | - | - | - |
| Atrial fibrillation and flutter | 25-29 | 0.000156 | 0.000003 | 0.000004 | - | - | 0.000081 | 0.000009 | 0.000010 | - | - |
| Atrial fibrillation and flutter | 30-34 | 0.000772 | 0.000023 | 0.000057 | - | - | 0.000411 | 0.000063 | 0.000157 | - | 0.101310 |
| Atrial fibrillation and flutter | 35-39 | 0.000570 | 0.000083 | 0.000302 | - | 0.095877 | 0.000399 | 0.000227 | 0.000830 | - | 0.094048 |
| Atrial fibrillation and flutter | 40-44 | 0.000417 | 0.000204 | 0.000995 | - | 0.097173 | 0.000378 | 0.000520 | 0.002646 | - | 0.095473 |
| Atrial fibrillation and flutter | 45-49 | 0.000558 | 0.000416 | 0.002500 | - | 0.097275 | 0.000436 | 0.000991 | 0.006324 | - | 0.095620 |
| Atrial fibrillation and flutter | 50-54 | 0.000582 | 0.000830 | 0.005457 | - | 0.097562 | 0.000441 | 0.001730 | 0.012902 | - | 0.095950 |
| Atrial fibrillation and flutter | 55-59 | 0.000667 | 0.001702 | 0.011423 | - | 0.098312 | 0.000544 | 0.002852 | 0.023914 | - | 0.096767 |
| Atrial fibrillation and flutter | 60-64 | 0.000907 | 0.003307 | 0.023359 | - | 0.098742 | 0.000753 | 0.004374 | 0.041176 | - | 0.097978 |
| Atrial fibrillation and flutter | 65-69 | 0.001193 | 0.005413 | 0.044164 | - | 0.099794 | 0.001030 | 0.005902 | 0.065372 | - | 0.099342 |
| Atrial fibrillation and flutter | 70-74 | 0.001895 | 0.006954 | 0.073207 | - | 0.101047 | 0.001637 | 0.006640 | 0.094113 | - | 0.101122 |

| | | | | | | | | | | | |
|---|---|---|---|---|---|---|---|---|---|---|---|
| Atrial fibrillation and flutter | 75-79 | 0.003450 | 0.007221 | 0.104832 | - | 0.102932 | 0.002894 | 0.006322 | 0.122198 | - | 0.102924 |
| Atrial fibrillation and flutter | 80-84 | 0.007231 | 0.006169 | 0.132137 | - | 0.105802 | 0.005820 | 0.005251 | 0.145058 | - | 0.105729 |
| Atrial fibrillation and flutter | 85-89 | 0.017576 | 0.004248 | 0.147568 | - | 0.109098 | 0.013097 | 0.003861 | 0.158550 | - | 0.108210 |
| Atrial fibrillation and flutter | 90-94 | 0.040176 | 0.002845 | 0.144738 | - | 0.113608 | 0.027574 | 0.002910 | 0.159221 | - | 0.110964 |
| Atrial fibrillation and flutter | 95+ | 0.065087 | 0.002306 | 0.124710 | - | 0.120468 | 0.044054 | 0.002579 | 0.147270 | - | 0.115853 |
| Breast cancer | 0-4 | - | - | - | - | - | | | | | |
| Breast cancer | 5-9 | 0.000268 | 0.000000 | 0.000000 | 0.000000 | - | | | | | |
| Breast cancer | 10-14 | 0.005633 | 0.000002 | 0.000005 | 0.000003 | - | | | | | |
| Breast cancer | 15-19 | 0.010705 | 0.000005 | 0.000024 | 0.000006 | 0.080212 | | | | | |
| Breast cancer | 20-24 | 0.013894 | 0.000012 | 0.000057 | 0.000006 | 0.100335 | | | | | |
| Breast cancer | 25-29 | 0.022769 | 0.000066 | 0.000191 | 0.000006 | 0.102260 | | | | | |
| Breast cancer | 30-34 | 0.025056 | 0.000263 | 0.000895 | 0.000006 | 0.100609 | | | | | |
| Breast cancer | 35-39 | 0.022924 | 0.000560 | 0.002708 | 0.000006 | 0.096704 | | | | | |
| Breast cancer | 40-44 | 0.019458 | 0.000985 | 0.006056 | 0.000006 | 0.094184 | | | | | |
| Breast cancer | 45-49 | 0.016898 | 0.001476 | 0.011405 | 0.000006 | 0.091080 | | | | | |
| Breast cancer | 50-54 | 0.016136 | 0.001912 | 0.018335 | 0.010186 | 0.089131 | | | | | |
| Breast cancer | 55-59 | 0.016409 | 0.002292 | 0.024321 | 0.033405 | 0.085509 | | | | | |
| Breast cancer | 60-64 | 0.016717 | 0.002767 | 0.030164 | 0.026383 | 0.083060 | | | | | |
| Breast cancer | 65-69 | 0.017308 | 0.003223 | 0.037500 | 0.028789 | 0.083100 | | | | | |
| Breast cancer | 70-74 | 0.018841 | 0.003474 | 0.043279 | 0.038336 | 0.083857 | | | | | |
| Breast cancer | 75-79 | 0.021912 | 0.003658 | 0.046789 | 0.040523 | 0.085840 | | | | | |

| | | | | | | | | | | | |
|---|---|---|---|---|---|---|---|---|---|---|---|
| Breast cancer | 80-84 | 0.027597 | 0.003904 | 0.049647 | 0.040361 | 0.092302 | | | | | |
| Breast cancer | 85-89 | 0.039022 | 0.003891 | 0.049400 | 0.050312 | 0.099444 | | | | | |
| Breast cancer | 90-94 | 0.060138 | 0.003320 | 0.043416 | 0.051231 | 0.093749 | | | | | |
| Breast cancer | 95+ | 0.082867 | 0.003520 | 0.037935 | 0.020102 | 0.084837 | | | | | |
| Cirrhosis | 0-4 | 0.000051 | 0.000119 | 0.004162 | - | 0.002120 | 0.000052 | 0.000151 | 0.004808 | - | 0.001185 |
| Cirrhosis | 5-9 | 0.000036 | 0.000844 | 0.006396 | - | 0.001150 | 0.000027 | 0.000872 | 0.007152 | - | 0.000762 |
| Cirrhosis | 10-14 | 0.000025 | 0.001551 | 0.012457 | - | 0.001041 | 0.000010 | 0.001978 | 0.014117 | - | 0.000661 |
| Cirrhosis | 15-19 | 0.000020 | 0.004787 | 0.026205 | - | 0.000371 | 0.000008 | 0.010418 | 0.039584 | - | 0.000136 |
| Cirrhosis | 20-24 | 0.000018 | 0.008974 | 0.061137 | - | 0.000228 | 0.000009 | 0.020863 | 0.117247 | - | 0.000078 |
| Cirrhosis | 25-29 | 0.000025 | 0.007039 | 0.100057 | - | 0.000138 | 0.000018 | 0.014996 | 0.197024 | - | 0.000050 |
| Cirrhosis | 30-34 | 0.000060 | 0.002097 | 0.120140 | - | 0.000136 | 0.000053 | 0.004218 | 0.233871 | - | 0.000057 |
| Cirrhosis | 35-39 | 0.000120 | 0.002042 | 0.127011 | - | 0.000173 | 0.000111 | 0.003434 | 0.244602 | - | 0.000078 |
| Cirrhosis | 40-44 | 0.000205 | 0.003008 | 0.138596 | - | 0.000250 | 0.000214 | 0.006227 | 0.263064 | - | 0.000162 |
| Cirrhosis | 45-49 | 0.000297 | 0.003760 | 0.152298 | - | 0.000289 | 0.000369 | 0.006235 | 0.286140 | - | 0.000243 |
| Cirrhosis | 50-54 | 0.000396 | 0.004108 | 0.169902 | - | 0.000367 | 0.000576 | 0.003727 | 0.303716 | - | 0.000344 |
| Cirrhosis | 55-59 | 0.000504 | 0.001765 | 0.181238 | - | 0.000421 | 0.000813 | 0.000670 | 0.310149 | - | 0.000430 |
| Cirrhosis | 60-64 | 0.000557 | 0.003231 | 0.189460 | - | 0.000399 | 0.000939 | 0.000469 | 0.310022 | - | 0.000416 |
| Cirrhosis | 65-69 | 0.000564 | 0.007690 | 0.210467 | - | 0.000334 | 0.000966 | 0.004014 | 0.316057 | - | 0.000351 |
| Cirrhosis | 70-74 | 0.000590 | 0.008183 | 0.242672 | - | 0.000353 | 0.000981 | 0.003243 | 0.329372 | - | 0.000369 |
| Cirrhosis | 75-79 | 0.000674 | 0.005367 | 0.267011 | - | 0.000345 | 0.001046 | 0.001796 | 0.334907 | - | 0.000342 |
| Cirrhosis | 80-84 | 0.000848 | 0.003012 | 0.281635 | - | 0.000324 | 0.001221 | 0.003168 | 0.342949 | - | 0.000345 |

| | | | | | | | | | | | |
|---|---|---|---|---|---|---|---|---|---|---|---|
| Cirrhosis | 85-89 | 0.001261 | 0.000538 | 0.286557 | - | 0.000326 | 0.001640 | 0.000660 | 0.347772 | - | 0.000341 |
| Cirrhosis | 90-94 | 0.002188 | 0.000006 | 0.285307 | - | 0.000413 | 0.002438 | 0.000006 | 0.346005 | - | 0.000396 |
| Cirrhosis | 95+ | 0.003487 | 0.000006 | 0.282455 | - | 0.000540 | 0.003239 | 0.000006 | 0.342804 | - | 0.000372 |
| Colon and rectum cancer | 0-4 | 0.010827 | 0.000001 | 0.000004 | 0.002884 | - | 0.010712 | 0.000002 | 0.000004 | 0.004543 | - |
| Colon and rectum cancer | 5-9 | 0.018404 | 0.000001 | 0.000009 | 0.053445 | 0.141345 | 0.019309 | 0.000002 | 0.000010 | 0.084253 | 0.139549 |
| Colon and rectum cancer | 10-14 | 0.024966 | 0.000002 | 0.000013 | 0.043168 | 0.129235 | 0.024431 | 0.000002 | 0.000012 | 0.068052 | 0.130250 |
| Colon and rectum cancer | 15-19 | 0.028131 | 0.000004 | 0.000023 | 0.002061 | 0.122211 | 0.028610 | 0.000004 | 0.000020 | 0.003246 | 0.123717 |
| Colon and rectum cancer | 20-24 | 0.031872 | 0.000011 | 0.000051 | 0.000006 | 0.125610 | 0.036902 | 0.000011 | 0.000049 | 0.000006 | 0.120000 |
| Colon and rectum cancer | 25-29 | 0.037398 | 0.000029 | 0.000129 | 0.000006 | 0.119658 | 0.041801 | 0.000030 | 0.000128 | 0.000006 | 0.116166 |
| Colon and rectum cancer | 30-34 | 0.037808 | 0.000067 | 0.000322 | 0.000006 | 0.110129 | 0.040324 | 0.000068 | 0.000322 | 0.000006 | 0.107443 |
| Colon and rectum cancer | 35-39 | 0.032753 | 0.000109 | 0.000673 | 0.000006 | 0.105032 | 0.034791 | 0.000115 | 0.000682 | 0.000006 | 0.106516 |
| Colon and rectum cancer | 40-44 | 0.030033 | 0.000179 | 0.001232 | 0.000006 | 0.104408 | 0.032539 | 0.000198 | 0.001287 | 0.000006 | 0.103462 |
| Colon and rectum cancer | 45-49 | 0.030906 | 0.000303 | 0.002163 | 0.000006 | 0.103780 | 0.034273 | 0.000355 | 0.002340 | 0.000006 | 0.100095 |
| Colon and rectum cancer | 50-54 | 0.031871 | 0.000479 | 0.003593 | 0.011198 | 0.097410 | 0.036052 | 0.000615 | 0.004134 | 0.002526 | 0.097879 |
| Colon and rectum cancer | 55-59 | 0.033397 | 0.000724 | 0.005229 | 0.037911 | 0.098768 | 0.037515 | 0.001036 | 0.006949 | 0.011597 | 0.100810 |
| Colon and rectum cancer | 60-64 | 0.037523 | 0.001120 | 0.007465 | 0.034264 | 0.101519 | 0.040990 | 0.001715 | 0.011099 | 0.024650 | 0.104345 |
| Colon and rectum cancer | 65-69 | 0.038829 | 0.001574 | 0.010949 | 0.029872 | 0.103497 | 0.043017 | 0.002526 | 0.016579 | 0.034059 | 0.106972 |
| Colon and rectum cancer | 70-74 | 0.041160 | 0.002229 | 0.015767 | 0.026145 | 0.109105 | 0.045433 | 0.003463 | 0.023189 | 0.040618 | 0.112642 |
| Colon and rectum cancer | 75-79 | 0.048625 | 0.003161 | 0.021824 | 0.041729 | 0.114033 | 0.053258 | 0.004554 | 0.029579 | 0.064612 | 0.122556 |
| Colon and rectum cancer | 80-84 | 0.065082 | 0.004146 | 0.025616 | 0.082642 | 0.127487 | 0.071390 | 0.005563 | 0.031711 | 0.103905 | 0.138392 |
| Colon and rectum cancer | 85-89 | 0.107076 | 0.004755 | 0.024416 | 0.122776 | 0.149893 | 0.110715 | 0.005970 | 0.029005 | 0.130182 | 0.162181 |

| | | | | | | | | | | | |
|---|---|---|---|---|---|---|---|---|---|---|---|
| Colon and rectum cancer | 90-94 | 0.205881 | 0.004765 | 0.017248 | 0.161918 | 0.185777 | 0.191419 | 0.005624 | 0.021137 | 0.154807 | 0.191002 |
| Colon and rectum cancer | 95+ | 0.331217 | 0.005047 | 0.012123 | 0.122125 | 0.241555 | 0.283209 | 0.005869 | 0.015598 | 0.121471 | 0.230758 |
| Diabetes mellitus type 2 | 0-4 | - | - | - | - | - | - | - | - | - | - |
| Diabetes mellitus type 2 | 5-9 | 0.000048 | 0.000001 | 0.000000 | - | - | 0.000071 | 0.000000 | 0.000000 | - | - |
| Diabetes mellitus type 2 | 10-14 | 0.001011 | 0.000011 | 0.000024 | - | - | 0.001496 | 0.000005 | 0.000010 | - | - |
| Diabetes mellitus type 2 | 15-19 | 0.001699 | 0.000030 | 0.000125 | - | - | 0.002584 | 0.000010 | 0.000049 | - | 0.055882 |
| Diabetes mellitus type 2 | 20-24 | 0.001056 | 0.000232 | 0.000596 | - | 0.059456 | 0.001677 | 0.000204 | 0.000390 | - | 0.058144 |
| Diabetes mellitus type 2 | 25-29 | 0.000359 | 0.000806 | 0.003132 | - | 0.062945 | 0.000489 | 0.000822 | 0.002845 | - | 0.059996 |
| Diabetes mellitus type 2 | 30-34 | 0.000251 | 0.001327 | 0.008453 | - | 0.067428 | 0.000323 | 0.001520 | 0.008638 | - | 0.064374 |
| Diabetes mellitus type 2 | 35-39 | 0.000234 | 0.001886 | 0.016337 | - | 0.073490 | 0.000323 | 0.002305 | 0.018008 | - | 0.071845 |
| Diabetes mellitus type 2 | 40-44 | 0.000250 | 0.002526 | 0.027073 | - | 0.079845 | 0.000331 | 0.003224 | 0.031401 | - | 0.078969 |
| Diabetes mellitus type 2 | 45-49 | 0.000347 | 0.003205 | 0.040864 | - | 0.086111 | 0.000514 | 0.004224 | 0.049188 | - | 0.086017 |
| Diabetes mellitus type 2 | 50-54 | 0.000472 | 0.003854 | 0.057572 | - | 0.092323 | 0.000807 | 0.005211 | 0.071180 | - | 0.092454 |
| Diabetes mellitus type 2 | 55-59 | 0.000606 | 0.004504 | 0.076833 | - | 0.098619 | 0.001036 | 0.006316 | 0.097028 | - | 0.098096 |
| Diabetes mellitus type 2 | 60-64 | 0.000850 | 0.005385 | 0.098998 | - | 0.105247 | 0.001390 | 0.008038 | 0.128063 | - | 0.104219 |
| Diabetes mellitus type 2 | 65-69 | 0.001146 | 0.006404 | 0.124662 | - | 0.112522 | 0.001864 | 0.009873 | 0.165420 | - | 0.110852 |
| Diabetes mellitus type 2 | 70-74 | 0.001647 | 0.007219 | 0.153267 | - | 0.117787 | 0.002546 | 0.009511 | 0.204214 | - | 0.115665 |
| Diabetes mellitus type 2 | 75-79 | 0.002701 | 0.007148 | 0.182049 | - | 0.120341 | 0.003800 | 0.006983 | 0.233938 | - | 0.119286 |
| Diabetes mellitus type 2 | 80-84 | 0.004858 | 0.005470 | 0.205271 | - | 0.123645 | 0.006223 | 0.004095 | 0.250345 | - | 0.123616 |
| Diabetes mellitus type 2 | 85-89 | 0.009170 | 0.002451 | 0.215226 | - | 0.128260 | 0.010604 | 0.001746 | 0.253227 | - | 0.129077 |
| Diabetes mellitus type 2 | 90-94 | 0.016931 | 0.000564 | 0.209908 | - | 0.135036 | 0.017280 | 0.000425 | 0.244013 | - | 0.135667 |

| | | | | | | | | | | | |
|---|---|---|---|---|---|---|---|---|---|---|---|
| Diabetes mellitus type 2 | 95+ | 0.026557 | 0.000073 | 0.193293 | - | 0.144814 | 0.023602 | 0.000078 | 0.226213 | - | 0.145183 |
| Epilepsy | 0-4 | 0.001317 | 0.000689 | 0.002265 | 0.300728 | 0.241891 | 0.001302 | 0.000730 | 0.002173 | 0.329412 | 0.242837 |
| Epilepsy | 5-9 | 0.000963 | 0.000592 | 0.002589 | 0.176979 | 0.242955 | 0.000965 | 0.000630 | 0.002511 | 0.197222 | 0.241937 |
| Epilepsy | 10-14 | 0.000780 | 0.000498 | 0.003370 | 0.101922 | 0.245122 | 0.000809 | 0.000547 | 0.003276 | 0.122469 | 0.243903 |
| Epilepsy | 15-19 | 0.001079 | 0.000434 | 0.003832 | 0.101503 | 0.248984 | 0.001231 | 0.000499 | 0.003702 | 0.121333 | 0.246688 |
| Epilepsy | 20-24 | 0.001643 | 0.000375 | 0.003815 | 0.106551 | 0.252916 | 0.002263 | 0.000444 | 0.003765 | 0.122413 | 0.250593 |
| Epilepsy | 25-29 | 0.002092 | 0.000326 | 0.003516 | 0.110375 | 0.254309 | 0.003630 | 0.000383 | 0.003399 | 0.138996 | 0.252508 |
| Epilepsy | 30-34 | 0.002493 | 0.000296 | 0.003190 | 0.107248 | 0.254936 | 0.004795 | 0.000346 | 0.002941 | 0.136467 | 0.252511 |
| Epilepsy | 35-39 | 0.002781 | 0.000286 | 0.002972 | 0.104086 | 0.255988 | 0.005750 | 0.000336 | 0.002708 | 0.127222 | 0.247774 |
| Epilepsy | 40-44 | 0.003028 | 0.000275 | 0.002854 | 0.098445 | 0.257303 | 0.006464 | 0.000323 | 0.002634 | 0.119415 | 0.253932 |
| Epilepsy | 45-49 | 0.003216 | 0.000260 | 0.002767 | 0.099727 | 0.257593 | 0.006874 | 0.000299 | 0.002569 | 0.115865 | 0.254741 |
| Epilepsy | 50-54 | 0.003175 | 0.000254 | 0.002625 | 0.099698 | 0.260058 | 0.006816 | 0.000278 | 0.002488 | 0.106966 | 0.256323 |
| Epilepsy | 55-59 | 0.002927 | 0.000259 | 0.002718 | 0.071170 | 0.258415 | 0.006095 | 0.000270 | 0.002590 | 0.078431 | 0.258035 |
| Epilepsy | 60-64 | 0.003062 | 0.000269 | 0.003179 | 0.042359 | 0.263797 | 0.005042 | 0.000280 | 0.003018 | 0.046964 | 0.260471 |
| Epilepsy | 65-69 | 0.003057 | 0.000284 | 0.003915 | 0.027210 | 0.260483 | 0.004351 | 0.000307 | 0.003804 | 0.026400 | 0.263103 |
| Epilepsy | 70-74 | 0.003330 | 0.000311 | 0.004743 | 0.028557 | 0.267714 | 0.004595 | 0.000339 | 0.004799 | 0.024952 | 0.264918 |
| Epilepsy | 75-79 | 0.004130 | 0.000347 | 0.005529 | 0.027409 | 0.270567 | 0.005669 | 0.000378 | 0.005689 | 0.031205 | 0.272214 |
| Epilepsy | 80-84 | 0.005612 | 0.000373 | 0.006476 | 0.023925 | 0.279111 | 0.007131 | 0.000446 | 0.006529 | 0.034205 | 0.278753 |
| Epilepsy | 85-89 | 0.007893 | 0.000390 | 0.007035 | 0.039754 | 0.289320 | 0.009054 | 0.000539 | 0.007432 | 0.037657 | 0.285100 |
| Epilepsy | 90-94 | 0.009720 | 0.000443 | 0.007316 | 0.039374 | 0.299904 | 0.011624 | 0.000588 | 0.008417 | 0.034056 | 0.292696 |
| Epilepsy | 95+ | 0.011134 | 0.000521 | 0.007779 | 0.043526 | 0.317099 | 0.011998 | 0.000573 | 0.009297 | 0.034041 | 0.302919 |

| | | | | | | | | | | | |
|---|---|---|---|---|---|---|---|---|---|---|---|
| Hypertensive heart disease | 0-4 | - | - | - | - | - | - | - | - | - | - |
| Hypertensive heart disease | 5-9 | 0.001209 | 0.000000 | 0.000000 | - | - | 0.001269 | 0.000000 | 0.000000 | - | - |
| Hypertensive heart disease | 10-14 | 0.025383 | 0.000000 | 0.000000 | - | - | 0.026652 | 0.000000 | 0.000000 | - | - |
| Hypertensive heart disease | 15-19 | 0.044251 | 0.000000 | 0.000001 | - | 0.101821 | 0.048157 | 0.000000 | 0.000001 | - | 0.100557 |
| Hypertensive heart disease | 20-24 | 0.031500 | 0.000001 | 0.000003 | - | 0.104787 | 0.041246 | 0.000001 | 0.000003 | - | 0.102547 |
| Hypertensive heart disease | 25-29 | 0.016454 | 0.000001 | 0.000007 | - | 0.106446 | 0.035303 | 0.000002 | 0.000010 | - | 0.103628 |
| Hypertensive heart disease | 30-34 | 0.014402 | 0.000004 | 0.000018 | - | 0.107162 | 0.033692 | 0.000005 | 0.000024 | - | 0.104380 |
| Hypertensive heart disease | 35-39 | 0.016289 | 0.000009 | 0.000044 | - | 0.108108 | 0.031309 | 0.000013 | 0.000059 | - | 0.105348 |
| Hypertensive heart disease | 40-44 | 0.014758 | 0.000021 | 0.000109 | - | 0.110934 | 0.026713 | 0.000035 | 0.000158 | - | 0.106194 |
| Hypertensive heart disease | 45-49 | 0.011034 | 0.000041 | 0.000251 | - | 0.102452 | 0.022296 | 0.000064 | 0.000377 | - | 0.102479 |
| Hypertensive heart disease | 50-54 | 0.009400 | 0.000045 | 0.000467 | - | 0.102097 | 0.021245 | 0.000065 | 0.000670 | - | 0.101715 |
| Hypertensive heart disease | 55-59 | 0.010835 | 0.000032 | 0.000607 | - | 0.103897 | 0.024368 | 0.000038 | 0.000811 | - | 0.103300 |
| Hypertensive heart disease | 60-64 | 0.013541 | 0.000138 | 0.000906 | - | 0.104317 | 0.027529 | 0.000153 | 0.001079 | - | 0.103915 |
| Hypertensive heart disease | 65-69 | 0.011316 | 0.000469 | 0.002259 | - | 0.105124 | 0.021798 | 0.000492 | 0.002423 | - | 0.106038 |
| Hypertensive heart disease | 70-74 | 0.009890 | 0.000652 | 0.004997 | - | 0.106938 | 0.016337 | 0.000622 | 0.005021 | - | 0.107485 |
| Hypertensive heart disease | 75-79 | 0.013165 | 0.000838 | 0.008198 | - | 0.109848 | 0.017650 | 0.000638 | 0.007548 | - | 0.110601 |
| Hypertensive heart disease | 80-84 | 0.021191 | 0.001536 | 0.012912 | - | 0.113209 | 0.024377 | 0.001022 | 0.010516 | - | 0.113132 |
| Hypertensive heart disease | 85-89 | 0.038173 | 0.002808 | 0.021097 | - | 0.117314 | 0.036499 | 0.001817 | 0.015575 | - | 0.116114 |
| Hypertensive heart disease | 90-94 | 0.072961 | 0.004109 | 0.030896 | - | 0.122268 | 0.058907 | 0.002496 | 0.021839 | - | 0.119404 |
| Hypertensive heart disease | 95+ | 0.119203 | 0.005608 | 0.038069 | - | 0.129469 | 0.092450 | 0.003252 | 0.026780 | - | 0.124669 |
| Intracerebral haemorrhage | 0-4 | 0.001903 | 0.000009 | 0.000064 | - | 0.160009 | 0.002690 | 0.000005 | 0.000036 | - | 0.128524 |

| | | | | | | | | | | | |
|---|---|---|---|---|---|---|---|---|---|---|---|
| Intracerebral haemorrhage | 5-9 | 0.001370 | 0.000020 | 0.000133 | - | 0.158294 | 0.001585 | 0.000012 | 0.000077 | - | 0.128114 |
| Intracerebral haemorrhage | 10-14 | 0.000980 | 0.000027 | 0.000251 | - | 0.162285 | 0.000946 | 0.000018 | 0.000154 | - | 0.131934 |
| Intracerebral haemorrhage | 15-19 | 0.001026 | 0.000027 | 0.000385 | - | 0.169805 | 0.001311 | 0.000019 | 0.000245 | - | 0.137692 |
| Intracerebral haemorrhage | 20-24 | 0.001310 | 0.000028 | 0.000520 | - | 0.177974 | 0.002202 | 0.000021 | 0.000342 | - | 0.143113 |
| Intracerebral haemorrhage | 25-29 | 0.001798 | 0.000031 | 0.000664 | - | 0.176468 | 0.003561 | 0.000024 | 0.000448 | - | 0.147594 |
| Intracerebral haemorrhage | 30-34 | 0.002969 | 0.000034 | 0.000818 | - | 0.177469 | 0.005942 | 0.000029 | 0.000567 | - | 0.149780 |
| Intracerebral haemorrhage | 35-39 | 0.004312 | 0.000036 | 0.000977 | - | 0.177289 | 0.008698 | 0.000036 | 0.000706 | - | 0.150639 |
| Intracerebral haemorrhage | 40-44 | 0.006503 | 0.000042 | 0.001141 | - | 0.176590 | 0.013618 | 0.000050 | 0.000875 | - | 0.147809 |
| Intracerebral haemorrhage | 45-49 | 0.009879 | 0.000052 | 0.001324 | - | 0.174898 | 0.020821 | 0.000071 | 0.001089 | - | 0.147585 |
| Intracerebral haemorrhage | 50-54 | 0.014355 | 0.000069 | 0.001539 | - | 0.170903 | 0.029265 | 0.000101 | 0.001362 | - | 0.150202 |
| Intracerebral haemorrhage | 55-59 | 0.020949 | 0.000093 | 0.001795 | - | 0.171709 | 0.040328 | 0.000138 | 0.001690 | - | 0.151226 |
| Intracerebral haemorrhage | 60-64 | 0.032568 | 0.000130 | 0.002091 | - | 0.177274 | 0.059017 | 0.000188 | 0.002042 | - | 0.154300 |
| Intracerebral haemorrhage | 65-69 | 0.051905 | 0.000179 | 0.002386 | - | 0.186411 | 0.094886 | 0.000252 | 0.002306 | - | 0.159049 |
| Intracerebral haemorrhage | 70-74 | 0.083582 | 0.000286 | 0.002673 | - | 0.206811 | 0.169668 | 0.000373 | 0.002330 | - | 0.180878 |
| Intracerebral haemorrhage | 75-79 | 0.160795 | 0.000502 | 0.002938 | - | 0.236173 | 0.344236 | 0.000622 | 0.002049 | - | 0.216862 |
| Intracerebral haemorrhage | 80-84 | 0.298766 | 0.000894 | 0.002973 | - | 0.263717 | 0.632955 | 0.001056 | 0.001719 | - | 0.251162 |
| Intracerebral haemorrhage | 85-89 | 0.500112 | 0.001553 | 0.003074 | - | 0.297676 | 1.019878 | 0.001721 | 0.001687 | - | 0.283218 |
| Intracerebral haemorrhage | 90-94 | 0.823965 | 0.002466 | 0.003023 | - | 0.331943 | 1.565053 | 0.002607 | 0.001665 | - | 0.310154 |
| Intracerebral haemorrhage | 95+ | 1.113975 | 0.003296 | 0.002962 | - | 0.368336 | 1.901718 | 0.003318 | 0.001731 | - | 0.350962 |
| Ischaemic heart disease | 0-4 | - | - | - | - | - | - | - | - | - | - |
| Ischaemic heart disease | 5-9 | 0.000046 | 0.000001 | 0.000001 | 0.000190 | - | 0.000088 | 0.000003 | 0.000001 | 0.007362 | - |

| | | | | | | | | | | | |
|---|---|---|---|---|---|---|---|---|---|---|---|
| Ischaemic heart disease | 10-14 | 0.000961 | 0.000022 | 0.000048 | 0.003980 | - | 0.001847 | 0.000061 | 0.000112 | 0.154606 | - |
| Ischaemic heart disease | 15-19 | 0.001823 | 0.000048 | 0.000220 | 0.029386 | 0.000793 | 0.003487 | 0.000133 | 0.000352 | 0.235116 | 0.004804 |
| Ischaemic heart disease | 20-24 | 0.001996 | 0.000088 | 0.000443 | 0.072318 | 0.010454 | 0.003769 | 0.000268 | 0.000783 | 0.142717 | 0.010224 |
| Ischaemic heart disease | 25-29 | 0.002795 | 0.000184 | 0.000846 | 0.086551 | 0.012037 | 0.005052 | 0.000647 | 0.002032 | 0.139293 | 0.008673 |
| Ischaemic heart disease | 30-34 | 0.004161 | 0.000262 | 0.001575 | 0.040803 | 0.011993 | 0.006718 | 0.001138 | 0.004450 | 0.101751 | 0.007970 |
| Ischaemic heart disease | 35-39 | 0.005559 | 0.000320 | 0.002757 | 0.008456 | 0.012847 | 0.007933 | 0.001708 | 0.008977 | 0.042198 | 0.008189 |
| Ischaemic heart disease | 40-44 | 0.006980 | 0.000497 | 0.004564 | 0.000006 | 0.016138 | 0.008333 | 0.002985 | 0.017979 | 0.021284 | 0.009375 |
| Ischaemic heart disease | 45-49 | 0.007738 | 0.000908 | 0.007692 | 0.004041 | 0.020055 | 0.008170 | 0.005526 | 0.033832 | 0.030083 | 0.010479 |
| Ischaemic heart disease | 50-54 | 0.007630 | 0.001637 | 0.012948 | 0.015441 | 0.021361 | 0.007882 | 0.009515 | 0.059537 | 0.039141 | 0.010163 |
| Ischaemic heart disease | 55-59 | 0.007711 | 0.002881 | 0.021140 | 0.029423 | 0.019481 | 0.007702 | 0.015123 | 0.094143 | 0.055544 | 0.009523 |
| Ischaemic heart disease | 60-64 | 0.008320 | 0.004912 | 0.034342 | 0.031335 | 0.021107 | 0.007960 | 0.022370 | 0.141486 | 0.047030 | 0.010729 |
| Ischaemic heart disease | 65-69 | 0.008982 | 0.007553 | 0.056988 | 0.012865 | 0.029068 | 0.008248 | 0.030515 | 0.211386 | 0.029800 | 0.014638 |
| Ischaemic heart disease | 70-74 | 0.010782 | 0.011149 | 0.092803 | 0.012663 | 0.036192 | 0.009304 | 0.040239 | 0.298663 | 0.029655 | 0.018673 |
| Ischaemic heart disease | 75-79 | 0.015863 | 0.015972 | 0.133900 | 0.026883 | 0.038470 | 0.012516 | 0.052380 | 0.378490 | 0.039320 | 0.020828 |
| Ischaemic heart disease | 80-84 | 0.028256 | 0.021664 | 0.175746 | 0.033168 | 0.042251 | 0.020197 | 0.067686 | 0.446800 | 0.045284 | 0.024137 |
| Ischaemic heart disease | 85-89 | 0.060356 | 0.030175 | 0.208751 | 0.042361 | 0.054551 | 0.037972 | 0.087611 | 0.493847 | 0.059393 | 0.030779 |
| Ischaemic heart disease | 90-94 | 0.133793 | 0.049466 | 0.225919 | 0.055335 | 0.070327 | 0.074377 | 0.113177 | 0.496048 | 0.084883 | 0.039012 |
| Ischaemic heart disease | 95+ | 0.227898 | 0.076930 | 0.242044 | 0.049524 | 0.080577 | 0.122290 | 0.141035 | 0.480148 | 0.091882 | 0.049331 |
| Ischaemic stroke | 0-4 | 0.000302 | 0.000007 | 0.000270 | - | 0.161765 | 0.000429 | 0.000004 | 0.000152 | - | 0.128426 |
| Ischaemic stroke | 5-9 | 0.000163 | 0.000080 | 0.000476 | - | 0.156636 | 0.000227 | 0.000045 | 0.000268 | - | 0.127531 |
| Ischaemic stroke | 10-14 | 0.000072 | 0.000105 | 0.000976 | - | 0.160744 | 0.000097 | 0.000059 | 0.000548 | - | 0.131224 |

| | | | | | | | | | | | |
|---|---|---|---|---|---|---|---|---|---|---|---|
| Ischaemic stroke | 15-19 | 0.000070 | 0.000086 | 0.001445 | - | 0.159386 | 0.000099 | 0.000050 | 0.000816 | - | 0.135602 |
| Ischaemic stroke | 20-24 | 0.000091 | 0.000089 | 0.001873 | - | 0.166588 | 0.000144 | 0.000053 | 0.001069 | - | 0.139753 |
| Ischaemic stroke | 25-29 | 0.000139 | 0.000110 | 0.002365 | - | 0.168886 | 0.000229 | 0.000062 | 0.001352 | - | 0.143246 |
| Ischaemic stroke | 30-34 | 0.000257 | 0.000140 | 0.002981 | - | 0.169871 | 0.000358 | 0.000084 | 0.001708 | - | 0.141057 |
| Ischaemic stroke | 35-39 | 0.000358 | 0.000187 | 0.003782 | - | 0.168402 | 0.000564 | 0.000126 | 0.002217 | - | 0.140167 |
| Ischaemic stroke | 40-44 | 0.000462 | 0.000261 | 0.004878 | - | 0.164243 | 0.000852 | 0.000204 | 0.003018 | - | 0.142667 |
| Ischaemic stroke | 45-49 | 0.000574 | 0.000355 | 0.006393 | - | 0.163506 | 0.001295 | 0.000305 | 0.004266 | - | 0.141067 |
| Ischaemic stroke | 50-54 | 0.000701 | 0.000482 | 0.008420 | - | 0.165060 | 0.001876 | 0.000440 | 0.006045 | - | 0.141781 |
| Ischaemic stroke | 55-59 | 0.001023 | 0.000680 | 0.011229 | - | 0.171137 | 0.002820 | 0.000659 | 0.008663 | - | 0.146747 |
| Ischaemic stroke | 60-64 | 0.001928 | 0.000963 | 0.015122 | - | 0.178393 | 0.005124 | 0.000950 | 0.012379 | - | 0.153200 |
| Ischaemic stroke | 65-69 | 0.003077 | 0.001486 | 0.020804 | - | 0.185359 | 0.007556 | 0.001470 | 0.017768 | - | 0.158057 |
| Ischaemic stroke | 70-74 | 0.006726 | 0.002245 | 0.029255 | - | 0.202994 | 0.013143 | 0.002129 | 0.025483 | - | 0.179582 |
| Ischaemic stroke | 75-79 | 0.015291 | 0.003225 | 0.040560 | - | 0.231949 | 0.024043 | 0.002866 | 0.034858 | - | 0.213264 |
| Ischaemic stroke | 80-84 | 0.036976 | 0.004558 | 0.053358 | - | 0.260825 | 0.051266 | 0.003668 | 0.043779 | - | 0.243267 |
| Ischaemic stroke | 85-89 | 0.099213 | 0.007078 | 0.062446 | - | 0.289659 | 0.133767 | 0.005215 | 0.045680 | - | 0.272361 |
| Ischaemic stroke | 90-94 | 0.245888 | 0.012891 | 0.059710 | - | 0.319928 | 0.330658 | 0.009796 | 0.036663 | - | 0.299198 |
| Ischaemic stroke | 95+ | 0.451657 | 0.022486 | 0.052057 | - | 0.355287 | 0.606623 | 0.016827 | 0.029009 | - | 0.330784 |
| Larynx and pharynx cancer | 0-4 | 0.008693 | 0.000001 | 0.000002 | - | - | 0.002043 | 0.000000 | 0.000001 | - | - |
| Larynx and pharynx cancer | 5-9 | 0.011543 | 0.000001 | 0.000007 | - | 0.072516 | 0.038953 | 0.000000 | 0.000003 | - | 0.081013 |
| Larynx and pharynx cancer | 10-14 | 0.012261 | 0.000001 | 0.000011 | - | 0.073312 | 0.054046 | 0.000000 | 0.000003 | - | 0.083460 |
| Larynx and pharynx cancer | 15-19 | 0.010665 | 0.000001 | 0.000015 | - | 0.076193 | 0.043055 | 0.000001 | 0.000005 | - | 0.089205 |

| | | | | | | | | | | | |
|---|---|---|---|---|---|---|---|---|---|---|---|
| Larynx and pharynx cancer | 20-24 | 0.010338 | 0.000002 | 0.000021 | - | 0.074131 | 0.033216 | 0.000002 | 0.000009 | - | 0.070456 |
| Larynx and pharynx cancer | 25-29 | 0.012393 | 0.000004 | 0.000032 | - | 0.077449 | 0.025800 | 0.000004 | 0.000020 | - | 0.073732 |
| Larynx and pharynx cancer | 30-34 | 0.014528 | 0.000009 | 0.000060 | - | 0.084777 | 0.026732 | 0.000007 | 0.000043 | - | 0.086337 |
| Larynx and pharynx cancer | 35-39 | 0.017161 | 0.000011 | 0.000105 | - | 0.090304 | 0.033645 | 0.000015 | 0.000087 | - | 0.096613 |
| Larynx and pharynx cancer | 40-44 | 0.023438 | 0.000016 | 0.000157 | - | 0.101025 | 0.044206 | 0.000035 | 0.000183 | - | 0.109598 |
| Larynx and pharynx cancer | 45-49 | 0.038245 | 0.000023 | 0.000224 | - | 0.107123 | 0.061231 | 0.000071 | 0.000372 | - | 0.117952 |
| Larynx and pharynx cancer | 50-54 | 0.057934 | 0.000031 | 0.000293 | - | 0.112389 | 0.084568 | 0.000123 | 0.000661 | - | 0.125751 |
| Larynx and pharynx cancer | 55-59 | 0.074400 | 0.000041 | 0.000364 | - | 0.123757 | 0.109829 | 0.000191 | 0.001032 | - | 0.128180 |
| Larynx and pharynx cancer | 60-64 | 0.096789 | 0.000051 | 0.000426 | - | 0.134218 | 0.138788 | 0.000252 | 0.001384 | - | 0.134286 |
| Larynx and pharynx cancer | 65-69 | 0.119079 | 0.000062 | 0.000467 | - | 0.136557 | 0.153017 | 0.000294 | 0.001637 | - | 0.136413 |
| Larynx and pharynx cancer | 70-74 | 0.137747 | 0.000072 | 0.000493 | - | 0.151021 | 0.161840 | 0.000332 | 0.001832 | - | 0.143113 |
| Larynx and pharynx cancer | 75-79 | 0.175402 | 0.000076 | 0.000486 | - | 0.153879 | 0.187783 | 0.000355 | 0.001926 | - | 0.144974 |
| Larynx and pharynx cancer | 80-84 | 0.210015 | 0.000081 | 0.000424 | - | 0.200289 | 0.222877 | 0.000359 | 0.001779 | - | 0.159929 |
| Larynx and pharynx cancer | 85-89 | 0.243841 | 0.000094 | 0.000420 | - | 0.210438 | 0.250221 | 0.000362 | 0.001616 | - | 0.174904 |
| Larynx and pharynx cancer | 90-94 | 0.503370 | 0.000085 | 0.000257 | - | 0.265149 | 0.322635 | 0.000309 | 0.001251 | - | 0.205240 |
| Larynx and pharynx cancer | 95+ | 0.752737 | 0.000081 | 0.000123 | - | 0.388872 | 0.360669 | 0.000265 | 0.000876 | - | 0.194414 |
| Lip and oral cavity cancer | 0-4 | 0.005473 | 0.000001 | 0.000002 | - | - | 0.005636 | 0.000001 | 0.000001 | - | - |
| Lip and oral cavity cancer | 5-9 | 0.021915 | 0.000001 | 0.000005 | - | 0.075081 | 0.023928 | 0.000001 | 0.000004 | - | 0.074730 |
| Lip and oral cavity cancer | 10-14 | 0.031163 | 0.000001 | 0.000009 | - | 0.077762 | 0.031973 | 0.000001 | 0.000006 | - | 0.076885 |
| Lip and oral cavity cancer | 15-19 | 0.028321 | 0.000002 | 0.000013 | - | 0.079842 | 0.028030 | 0.000001 | 0.000010 | - | 0.079564 |
| Lip and oral cavity cancer | 20-24 | 0.026543 | 0.000003 | 0.000022 | - | 0.083504 | 0.027633 | 0.000003 | 0.000019 | - | 0.083715 |

| | | | | | | | | | | | |
|---|---|---|---|---|---|---|---|---|---|---|---|
| Lip and oral cavity cancer | 25-29 | 0.033336 | 0.000006 | 0.000039 | - | 0.086007 | 0.032478 | 0.000007 | 0.000040 | - | 0.084048 |
| Lip and oral cavity cancer | 30-34 | 0.042463 | 0.000008 | 0.000064 | - | 0.088647 | 0.037681 | 0.000015 | 0.000081 | - | 0.087612 |
| Lip and oral cavity cancer | 35-39 | 0.050500 | 0.000011 | 0.000094 | - | 0.090033 | 0.045524 | 0.000025 | 0.000156 | - | 0.088957 |
| Lip and oral cavity cancer | 40-44 | 0.068796 | 0.000016 | 0.000127 | - | 0.097320 | 0.063295 | 0.000038 | 0.000258 | - | 0.095619 |
| Lip and oral cavity cancer | 45-49 | 0.119063 | 0.000023 | 0.000156 | - | 0.105517 | 0.082571 | 0.000058 | 0.000375 | - | 0.103744 |
| Lip and oral cavity cancer | 50-54 | 0.198864 | 0.000030 | 0.000160 | - | 0.114033 | 0.100242 | 0.000093 | 0.000542 | - | 0.110617 |
| Lip and oral cavity cancer | 55-59 | 0.290693 | 0.000038 | 0.000146 | - | 0.134689 | 0.135719 | 0.000136 | 0.000740 | - | 0.114245 |
| Lip and oral cavity cancer | 60-64 | 0.406337 | 0.000049 | 0.000126 | - | 0.169776 | 0.190705 | 0.000177 | 0.000857 | - | 0.129891 |
| Lip and oral cavity cancer | 65-69 | 0.427210 | 0.000068 | 0.000141 | - | 0.188006 | 0.203573 | 0.000230 | 0.000962 | - | 0.140113 |
| Lip and oral cavity cancer | 70-74 | 0.424666 | 0.000093 | 0.000192 | - | 0.201506 | 0.204122 | 0.000277 | 0.001161 | - | 0.136846 |
| Lip and oral cavity cancer | 75-79 | 0.482928 | 0.000122 | 0.000239 | - | 0.209262 | 0.268651 | 0.000311 | 0.001233 | - | 0.148565 |
| Lip and oral cavity cancer | 80-84 | 0.614660 | 0.000164 | 0.000261 | - | 0.268790 | 0.431377 | 0.000342 | 0.000963 | - | 0.189621 |
| Lip and oral cavity cancer | 85-89 | 0.874617 | 0.000219 | 0.000260 | - | 0.302463 | 0.743718 | 0.000378 | 0.000612 | - | 0.267115 |
| Lip and oral cavity cancer | 90-94 | 1.417947 | 0.000271 | 0.000201 | - | 0.428395 | 1.364101 | 0.000387 | 0.000315 | - | 0.395132 |
| Lip and oral cavity cancer | 95+ | 1.847232 | 0.000277 | 0.000155 | - | 0.602575 | 1.836414 | 0.000360 | 0.000204 | - | 0.620887 |
| Liver cancer | 0-4 | 0.422951 | 0.000001 | 0.000002 | - | 0.176383 | 0.323901 | 0.000001 | 0.000002 | - | 0.107648 |
| Liver cancer | 5-9 | 0.211512 | 0.000001 | 0.000003 | - | 0.099828 | 0.173886 | 0.000001 | 0.000004 | - | 0.100325 |
| Liver cancer | 10-14 | 0.106864 | 0.000001 | 0.000006 | - | 0.102173 | 0.096446 | 0.000001 | 0.000007 | - | 0.105115 |
| Liver cancer | 15-19 | 0.133118 | 0.000000 | 0.000005 | - | 0.111714 | 0.112776 | 0.000000 | 0.000006 | - | 0.110453 |
| Liver cancer | 20-24 | 0.176929 | 0.000001 | 0.000004 | - | 0.136662 | 0.134781 | 0.000001 | 0.000007 | - | 0.124130 |
| Liver cancer | 25-29 | 0.280096 | 0.000001 | 0.000005 | - | 0.142884 | 0.180978 | 0.000003 | 0.000011 | - | 0.121941 |

| | | | | | | | | | | | |
|---|---|---|---|---|---|---|---|---|---|---|---|
| Liver cancer | 30-34 | 0.376228 | 0.000002 | 0.000006 | - | 0.163073 | 0.242806 | 0.000006 | 0.000018 | - | 0.134746 |
| Liver cancer | 35-39 | 0.418575 | 0.000004 | 0.000008 | - | 0.171527 | 0.268777 | 0.000011 | 0.000028 | - | 0.140303 |
| Liver cancer | 40-44 | 0.434064 | 0.000008 | 0.000014 | - | 0.176748 | 0.277937 | 0.000022 | 0.000053 | - | 0.143082 |
| Liver cancer | 45-49 | 0.469894 | 0.000016 | 0.000027 | - | 0.189454 | 0.300166 | 0.000047 | 0.000108 | - | 0.155189 |
| Liver cancer | 50-54 | 0.533077 | 0.000030 | 0.000047 | - | 0.198609 | 0.353246 | 0.000099 | 0.000207 | - | 0.168899 |
| Liver cancer | 55-59 | 0.609002 | 0.000051 | 0.000075 | - | 0.212472 | 0.442348 | 0.000174 | 0.000343 | - | 0.186217 |
| Liver cancer | 60-64 | 0.739590 | 0.000072 | 0.000092 | - | 0.233214 | 0.536614 | 0.000214 | 0.000395 | - | 0.199665 |
| Liver cancer | 65-69 | 0.856677 | 0.000107 | 0.000116 | - | 0.259810 | 0.635206 | 0.000273 | 0.000417 | - | 0.222447 |
| Liver cancer | 70-74 | 0.949224 | 0.000162 | 0.000159 | - | 0.277141 | 0.760003 | 0.000362 | 0.000463 | - | 0.260062 |
| Liver cancer | 75-79 | 1.082298 | 0.000230 | 0.000205 | - | 0.315212 | 0.905603 | 0.000457 | 0.000499 | - | 0.291792 |
| Liver cancer | 80-84 | 1.277121 | 0.000295 | 0.000229 | - | 0.354783 | 1.047324 | 0.000535 | 0.000511 | - | 0.328317 |
| Liver cancer | 85-89 | 1.541505 | 0.000322 | 0.000216 | - | 0.411713 | 1.288314 | 0.000551 | 0.000457 | - | 0.362039 |
| Liver cancer | 90-94 | 2.060979 | 0.000288 | 0.000148 | - | 0.515738 | 1.928356 | 0.000424 | 0.000244 | - | 0.502018 |
| Liver cancer | 95+ | 2.568673 | 0.000270 | 0.000107 | - | 0.684875 | 2.481620 | 0.000337 | 0.000141 | - | 0.667456 |
| Oesophageal cancer | 0-4 | - | - | - | - | - | - | - | - | - | - |
| Oesophageal cancer | 5-9 | - | - | - | - | - | - | - | - | - | - |
| Oesophageal cancer | 10-14 | - | - | - | - | - | - | - | - | - | - |
| Oesophageal cancer | 15-19 | 0.034096 | 0.000000 | 0.000000 | - | - | 0.067350 | 0.000000 | 0.000000 | - | - |
| Oesophageal cancer | 20-24 | 0.085178 | 0.000000 | 0.000001 | - | 0.112829 | 0.172028 | 0.000001 | 0.000002 | - | 0.129145 |
| Oesophageal cancer | 25-29 | 0.091585 | 0.000001 | 0.000003 | - | 0.118527 | 0.207007 | 0.000001 | 0.000004 | - | 0.140224 |
| Oesophageal cancer | 30-34 | 0.128623 | 0.000001 | 0.000005 | - | 0.124925 | 0.338569 | 0.000003 | 0.000006 | - | 0.156967 |

| | | | | | | | | | | | |
|---|---|---|---|---|---|---|---|---|---|---|---|
| Oesophageal cancer | 35-39 | 0.156919 | 0.000003 | 0.000009 | - | 0.130106 | 0.431107 | 0.000007 | 0.000012 | - | 0.158534 |
| Oesophageal cancer | 40-44 | 0.158454 | 0.000006 | 0.000020 | - | 0.127194 | 0.449443 | 0.000019 | 0.000030 | - | 0.175073 |
| Oesophageal cancer | 45-49 | 0.149295 | 0.000013 | 0.000043 | - | 0.122109 | 0.407236 | 0.000045 | 0.000077 | - | 0.168358 |
| Oesophageal cancer | 50-54 | 0.151796 | 0.000024 | 0.000088 | - | 0.137312 | 0.361124 | 0.000090 | 0.000176 | - | 0.176908 |
| Oesophageal cancer | 55-59 | 0.172753 | 0.000034 | 0.000140 | - | 0.149286 | 0.393706 | 0.000150 | 0.000314 | - | 0.186577 |
| Oesophageal cancer | 60-64 | 0.208830 | 0.000048 | 0.000185 | - | 0.152544 | 0.488258 | 0.000222 | 0.000419 | - | 0.203407 |
| Oesophageal cancer | 65-69 | 0.253995 | 0.000085 | 0.000256 | - | 0.160182 | 0.553189 | 0.000316 | 0.000522 | - | 0.203305 |
| Oesophageal cancer | 70-74 | 0.277333 | 0.000143 | 0.000391 | - | 0.176526 | 0.595499 | 0.000432 | 0.000670 | - | 0.226035 |
| Oesophageal cancer | 75-79 | 0.284035 | 0.000221 | 0.000603 | - | 0.207505 | 0.638708 | 0.000554 | 0.000822 | - | 0.249654 |
| Oesophageal cancer | 80-84 | 0.311294 | 0.000319 | 0.000865 | - | 0.190116 | 0.702420 | 0.000675 | 0.000936 | - | 0.276084 |
| Oesophageal cancer | 85-89 | 0.423580 | 0.000404 | 0.000999 | - | 0.249667 | 0.856926 | 0.000768 | 0.000937 | - | 0.316282 |
| Oesophageal cancer | 90-94 | 0.782550 | 0.000423 | 0.000669 | - | 0.346632 | 1.233367 | 0.000728 | 0.000649 | - | 0.401529 |
| Oesophageal cancer | 95+ | 1.570399 | 0.000481 | 0.000332 | - | 0.545001 | 1.750744 | 0.000705 | 0.000422 | - | 0.603865 |
| Pancreatitis | 0-4 | 0.000861 | 0.000034 | 0.000029 | 1.098036 | 0.124014 | 0.002171 | 0.000018 | 0.000013 | 1.313341 | 0.119795 |
| Pancreatitis | 5-9 | 0.000741 | 0.000048 | 0.000049 | 0.831822 | 0.091677 | 0.001233 | 0.000027 | 0.000022 | 1.082296 | 0.102970 |
| Pancreatitis | 10-14 | 0.000576 | 0.000078 | 0.000093 | 0.736849 | 0.093346 | 0.000783 | 0.000045 | 0.000041 | 1.005662 | 0.107181 |
| Pancreatitis | 15-19 | 0.000470 | 0.000150 | 0.000148 | 0.917696 | 0.114950 | 0.001413 | 0.000084 | 0.000066 | 1.163309 | 0.125558 |
| Pancreatitis | 20-24 | 0.000702 | 0.000264 | 0.000218 | 1.143835 | 0.135763 | 0.002929 | 0.000151 | 0.000104 | 1.341284 | 0.136442 |
| Pancreatitis | 25-29 | 0.001513 | 0.000369 | 0.000285 | 1.240521 | 0.142640 | 0.005400 | 0.000243 | 0.000160 | 1.413978 | 0.138573 |
| Pancreatitis | 30-34 | 0.002422 | 0.000415 | 0.000342 | 1.174796 | 0.135994 | 0.008590 | 0.000353 | 0.000233 | 1.425412 | 0.135855 |
| Pancreatitis | 35-39 | 0.003042 | 0.000410 | 0.000395 | 1.003637 | 0.123458 | 0.010211 | 0.000440 | 0.000312 | 1.337203 | 0.131119 |

| | | | | | | | | | | | |
|---|---|---|---|---|---|---|---|---|---|---|---|
| Pancreatitis | 40-44 | 0.004177 | 0.000402 | 0.000461 | 0.830734 | 0.108594 | 0.011513 | 0.000509 | 0.000404 | 1.191723 | 0.120698 |
| Pancreatitis | 45-49 | 0.005411 | 0.000419 | 0.000551 | 0.718601 | 0.103879 | 0.012911 | 0.000569 | 0.000514 | 1.046011 | 0.115445 |
| Pancreatitis | 50-54 | 0.006037 | 0.000451 | 0.000651 | 0.651669 | 0.099410 | 0.013867 | 0.000624 | 0.000634 | 0.927128 | 0.106993 |
| Pancreatitis | 55-59 | 0.006695 | 0.000492 | 0.000771 | 0.594985 | 0.096624 | 0.014740 | 0.000697 | 0.000776 | 0.836580 | 0.104184 |
| Pancreatitis | 60-64 | 0.008186 | 0.000571 | 0.000914 | 0.581385 | 0.096984 | 0.015788 | 0.000825 | 0.000982 | 0.772671 | 0.102282 |
| Pancreatitis | 65-69 | 0.010646 | 0.000691 | 0.001083 | 0.592004 | 0.094595 | 0.017093 | 0.001012 | 0.001227 | 0.766178 | 0.099647 |
| Pancreatitis | 70-74 | 0.015183 | 0.000849 | 0.001290 | 0.605301 | 0.092811 | 0.021543 | 0.001230 | 0.001431 | 0.811743 | 0.104749 |
| Pancreatitis | 75-79 | 0.025831 | 0.001058 | 0.001502 | 0.653747 | 0.098923 | 0.031784 | 0.001440 | 0.001540 | 0.893450 | 0.101110 |
| Pancreatitis | 80-84 | 0.045568 | 0.001316 | 0.001638 | 0.740206 | 0.112441 | 0.051406 | 0.001614 | 0.001542 | 1.000539 | 0.111809 |
| Pancreatitis | 85-89 | 0.077703 | 0.001581 | 0.001775 | 0.794744 | 0.108132 | 0.084422 | 0.001752 | 0.001459 | 1.132305 | 0.124685 |
| Pancreatitis | 90-94 | 0.129020 | 0.001792 | 0.001905 | 0.796587 | 0.108975 | 0.125295 | 0.001864 | 0.001343 | 1.278098 | 0.150821 |
| Pancreatitis | 95+ | 0.171612 | 0.001917 | 0.002007 | 0.773382 | 0.112242 | 0.162785 | 0.001927 | 0.001276 | 1.351917 | 0.160506 |

*The majority of diabetes-related deaths are from IHD and stroke

We derived incidence, prevalence and case fatality from Global Burden of Disease data[3] using DisMod II software (SF Figure 1)[5] to estimate the epidemiological inputs to the model.

#### Disease inputs- Acute causes

*Table S3: Epidemiological data for acute causes (rates per 1)*

| Cause | Age group (years) | Female | | | Male | | |
|---|---|---|---|---|---|---|---|
| | | Incidence | Mortality | YLD (disability) | Incidence | Mortality | YLD (disability) |
| Alcohol use disorders | 0-4 | - | - | 0.004643 | - | - | 0.002034 |
| Alcohol use disorders | 5-9 | 0.000406 | - | 0.040789 | - | - | 0.038766 |
| Alcohol use disorders | 10-14 | 0.002700 | 0.000000 | 0.090876 | 0.003235 | 0.000000 | 0.085186 |
| Alcohol use disorders | 15-19 | 0.007075 | 0.000001 | 0.163541 | 0.010126 | 0.000002 | 0.148813 |
| Alcohol use disorders | 20-24 | 0.010506 | 0.000002 | 0.202010 | 0.017621 | 0.000008 | 0.181153 |
| Alcohol use disorders | 25-29 | 0.011989 | 0.000005 | 0.206401 | 0.023109 | 0.000015 | 0.188015 |
| Alcohol use disorders | 30-34 | 0.011885 | 0.000008 | 0.194303 | 0.025801 | 0.000026 | 0.184639 |
| Alcohol use disorders | 35-39 | 0.011542 | 0.000013 | 0.185404 | 0.025955 | 0.000037 | 0.186877 |
| Alcohol use disorders | 40-44 | 0.010820 | 0.000018 | 0.194848 | 0.023155 | 0.000050 | 0.204427 |
| Alcohol use disorders | 45-49 | 0.009271 | 0.000021 | 0.211596 | 0.018221 | 0.000057 | 0.222313 |
| Alcohol use disorders | 50-54 | 0.007390 | 0.000020 | 0.225966 | 0.013543 | 0.000058 | 0.231306 |
| Alcohol use disorders | 55-59 | 0.005706 | 0.000020 | 0.239938 | 0.010667 | 0.000058 | 0.234574 |
| Alcohol use disorders | 60-64 | 0.004311 | 0.000020 | 0.254388 | 0.009040 | 0.000064 | 0.239558 |
| Alcohol use disorders | 65-69 | 0.003242 | 0.000020 | 0.262769 | 0.007611 | 0.000072 | 0.248130 |
| Alcohol use disorders | 70-74 | 0.002621 | 0.000019 | 0.261533 | 0.006335 | 0.000079 | 0.256354 |
| Alcohol use disorders | 75-79 | 0.002370 | 0.000020 | 0.258331 | 0.005395 | 0.000084 | 0.264566 |
| Alcohol use disorders | 80-84 | 0.002414 | 0.000023 | 0.254654 | 0.004813 | 0.000090 | 0.272536 |
| Alcohol use disorders | 85-89 | 0.003107 | 0.000028 | 0.251424 | 0.004709 | 0.000092 | 0.280264 |
| Alcohol use disorders | 90-94 | 0.004547 | 0.000034 | 0.250771 | 0.005103 | 0.000089 | 0.289218 |
| Alcohol use disorders | 95+ | 0.006738 | 0.000041 | 0.251279 | 0.005994 | 0.000082 | 0.298957 |
| Interpersonal violence | 0-4 | 0.003040 | 0.000008 | 0.016635 | 0.002707 | 0.000011 | 0.013075 |
| Interpersonal violence | 5-9 | 0.003829 | 0.000003 | 0.036871 | 0.006935 | 0.000004 | 0.017253 |
| Interpersonal violence | 10-14 | 0.005449 | 0.000002 | 0.052490 | 0.014009 | 0.000004 | 0.020195 |
| Interpersonal violence | 15-19 | 0.008227 | 0.000006 | 0.062702 | 0.025303 | 0.000013 | 0.020642 |
| Interpersonal violence | 20-24 | 0.009617 | 0.000010 | 0.065547 | 0.030632 | 0.000022 | 0.026328 |
| Interpersonal violence | 25-29 | 0.009242 | 0.000012 | 0.069746 | 0.027857 | 0.000028 | 0.041320 |
| Interpersonal violence | 30-34 | 0.007539 | 0.000013 | 0.084505 | 0.019973 | 0.000029 | 0.065745 |
| Interpersonal violence | 35-39 | 0.005617 | 0.000012 | 0.119825 | 0.013474 | 0.000029 | 0.097777 |
| Interpersonal violence | 40-44 | 0.003992 | 0.000011 | 0.178876 | 0.010001 | 0.000027 | 0.139869 |
| Interpersonal violence | 45-49 | 0.002766 | 0.000010 | 0.263357 | 0.007624 | 0.000024 | 0.204360 |
| Interpersonal violence | 50-54 | 0.001938 | 0.000009 | 0.371729 | 0.005532 | 0.000021 | 0.303156 |
| Interpersonal violence | 55-59 | 0.001418 | 0.000007 | 0.494414 | 0.003876 | 0.000018 | 0.436767 |
| Interpersonal violence | 60-64 | 0.001119 | 0.000007 | 0.609881 | 0.002811 | 0.000015 | 0.582167 |
| Interpersonal violence | 65-69 | 0.000977 | 0.000007 | 0.690053 | 0.002247 | 0.000014 | 0.707298 |
| Interpersonal violence | 70-74 | 0.000946 | 0.000007 | 0.707591 | 0.001956 | 0.000013 | 0.789463 |
| Interpersonal violence | 75-79 | 0.001038 | 0.000008 | 0.648526 | 0.001785 | 0.000012 | 0.807922 |
| Interpersonal violence | 80-84 | 0.001251 | 0.000007 | 0.534774 | 0.001800 | 0.000010 | 0.736337 |
| Interpersonal violence | 85-89 | 0.001463 | 0.000007 | 0.432937 | 0.002389 | 0.000010 | 0.600539 |
| Interpersonal violence | 90-94 | 0.001655 | 0.000007 | 0.345144 | 0.003585 | 0.000017 | 0.420097 |
| Interpersonal violence | 95+ | 0.001826 | 0.000009 | 0.272527 | 0.005393 | 0.000029 | 0.192357 |
| Lower respiratory infections | 0-4 | 0.028481 | 0.000029 | 0.001601 | 0.040632 | 0.000035 | 0.001629 |
| Lower respiratory infections | 5-9 | 0.015942 | 0.000007 | 0.001483 | 0.020342 | 0.000008 | 0.001496 |
| Lower respiratory infections | 10-14 | 0.008151 | - | 0.001416 | 0.008358 | - | 0.001421 |
| Lower respiratory infections | 15-19 | 0.006355 | 0.000002 | 0.001386 | 0.006256 | 0.000001 | 0.001389 |
| Lower respiratory infections | 20-24 | 0.007206 | 0.000002 | 0.001357 | 0.007022 | 0.000002 | 0.001361 |
| Lower respiratory infections | 25-29 | 0.008455 | 0.000003 | 0.001345 | 0.008313 | 0.000004 | 0.001346 |
| Lower respiratory infections | 30-34 | 0.009733 | 0.000005 | 0.001336 | 0.009762 | 0.000006 | 0.001338 |
| Lower respiratory infections | 35-39 | 0.011184 | 0.000006 | 0.001324 | 0.011708 | 0.000010 | 0.001328 |

| | | | | | | | |
|---|---|---|---|---|---|---|---|
| Lower respiratory infections | 40-44 | 0.012926 | 0.000009 | 0.001311 | 0.014549 | 0.000015 | 0.001317 |
| Lower respiratory infections | 45-49 | 0.014804 | 0.000012 | 0.001305 | 0.018091 | 0.000021 | 0.001315 |
| Lower respiratory infections | 50-54 | 0.016850 | 0.000018 | 0.001305 | 0.022124 | 0.000028 | 0.001305 |
| Lower respiratory infections | 55-59 | 0.020148 | 0.000025 | 0.001292 | 0.027807 | 0.000040 | 0.001277 |
| Lower respiratory infections | 60-64 | 0.025851 | 0.000035 | 0.001255 | 0.036621 | 0.000061 | 0.001237 |
| Lower respiratory infections | 65-69 | 0.032007 | 0.000047 | 0.001210 | 0.046239 | 0.000092 | 0.001196 |
| Lower respiratory infections | 70-74 | 0.039351 | 0.000081 | 0.001162 | 0.057433 | 0.000168 | 0.001158 |
| Lower respiratory infections | 75-79 | 0.058788 | 0.000254 | 0.001114 | 0.083312 | 0.000406 | 0.001116 |
| Lower respiratory infections | 80-84 | 0.101460 | 0.000689 | 0.001082 | 0.137365 | 0.001027 | 0.001088 |
| Lower respiratory infections | 85-89 | 0.163112 | 0.003301 | 0.001074 | 0.214394 | 0.003891 | 0.001086 |
| Lower respiratory infections | 90-94 | 0.238852 | 0.008519 | 0.001086 | 0.308459 | 0.009350 | 0.001105 |
| Lower respiratory infections | 95+ | 0.328636 | 0.016290 | 0.001119 | 0.419505 | 0.017365 | 0.001144 |
| Self-harm | 0-4 | - | - | - | - | - | - |
| Self-harm | 5-9 | 0.000433 | - | 0.002152 | 0.000142 | - | 0.003454 |
| Self-harm | 10-14 | 0.001128 | 0.000010 | 0.011533 | 0.000610 | 0.000007 | 0.011702 |
| Self-harm | 15-19 | 0.002169 | 0.000042 | 0.027385 | 0.001449 | 0.000134 | 0.024344 |
| Self-harm | 20-24 | 0.002429 | 0.000061 | 0.056644 | 0.001904 | 0.000217 | 0.047484 |
| Self-harm | 25-29 | 0.002146 | 0.000065 | 0.094142 | 0.001918 | 0.000257 | 0.078000 |
| Self-harm | 30-34 | 0.001685 | 0.000068 | 0.133550 | 0.001611 | 0.000255 | 0.117174 |
| Self-harm | 35-39 | 0.001492 | 0.000074 | 0.167519 | 0.001373 | 0.000260 | 0.160228 |
| Self-harm | 40-44 | 0.001410 | 0.000077 | 0.202768 | 0.001198 | 0.000256 | 0.216880 |
| Self-harm | 45-49 | 0.001222 | 0.000081 | 0.254959 | 0.000958 | 0.000253 | 0.300753 |
| Self-harm | 50-54 | 0.000948 | 0.000081 | 0.381098 | 0.000728 | 0.000239 | 0.428536 |
| Self-harm | 55-59 | 0.000648 | 0.000072 | 0.639303 | 0.000522 | 0.000221 | 0.648072 |
| Self-harm | 60-64 | 0.000400 | 0.000056 | 1.038831 | 0.000348 | 0.000192 | 1.007551 |
| Self-harm | 65-69 | 0.000254 | 0.000051 | 1.443487 | 0.000232 | 0.000173 | 1.398636 |
| Self-harm | 70-74 | 0.000220 | 0.000054 | 1.587322 | 0.000192 | 0.000177 | 1.541457 |
| Self-harm | 75-79 | 0.000253 | 0.000059 | 1.419436 | 0.000245 | 0.000202 | 1.312930 |
| Self-harm | 80-84 | 0.000289 | 0.000061 | 1.136990 | 0.000337 | 0.000259 | 0.894287 |
| Self-harm | 85-89 | 0.000281 | 0.000061 | 1.061302 | 0.000361 | 0.000325 | 0.730336 |
| Self-harm | 90-94 | 0.000238 | 0.000057 | 1.164753 | 0.000319 | 0.000355 | 0.805482 |
| Self-harm | 95+ | 0.000160 | 0.000050 | 1.451724 | 0.000209 | 0.000355 | 1.125497 |
| Transport injuries | 0-4 | 0.001839 | 0.000010 | 0.013794 | 0.001718 | 0.000012 | 0.013999 |
| Transport injuries | 5-9 | 0.004499 | 0.000015 | 0.018370 | 0.004796 | 0.000021 | 0.018312 |
| Transport injuries | 10-14 | 0.006726 | 0.000024 | 0.026396 | 0.007608 | 0.000050 | 0.025782 |
| Transport injuries | 15-19 | 0.008698 | 0.000040 | 0.036922 | 0.010402 | 0.000107 | 0.035447 |
| Transport injuries | 20-24 | 0.008885 | 0.000045 | 0.057773 | 0.010992 | 0.000146 | 0.055615 |
| Transport injuries | 25-29 | 0.007850 | 0.000040 | 0.089749 | 0.009880 | 0.000146 | 0.087550 |
| Transport injuries | 30-34 | 0.006477 | 0.000030 | 0.128972 | 0.008317 | 0.000123 | 0.123252 |
| Transport injuries | 35-39 | 0.005679 | 0.000026 | 0.168216 | 0.007627 | 0.000109 | 0.154768 |
| Transport injuries | 40-44 | 0.005326 | 0.000026 | 0.207603 | 0.007379 | 0.000106 | 0.190243 |
| Transport injuries | 45-49 | 0.004976 | 0.000028 | 0.256371 | 0.006818 | 0.000105 | 0.244268 |
| Transport injuries | 50-54 | 0.004477 | 0.000031 | 0.324996 | 0.005834 | 0.000100 | 0.325426 |
| Transport injuries | 55-59 | 0.003876 | 0.000034 | 0.418603 | 0.004796 | 0.000096 | 0.429874 |
| Transport injuries | 60-64 | 0.003306 | 0.000038 | 0.532923 | 0.004015 | 0.000099 | 0.550971 |
| Transport injuries | 65-69 | 0.002934 | 0.000046 | 0.634583 | 0.003427 | 0.000110 | 0.679174 |
| Transport injuries | 70-74 | 0.002869 | 0.000061 | 0.684079 | 0.003063 | 0.000134 | 0.772541 |
| Transport injuries | 75-79 | 0.003033 | 0.000091 | 0.680424 | 0.003073 | 0.000182 | 0.781148 |
| Transport injuries | 80-84 | 0.003219 | 0.000124 | 0.653164 | 0.003489 | 0.000256 | 0.701590 |
| Transport injuries | 85-89 | 0.003256 | 0.000146 | 0.639013 | 0.004155 | 0.000326 | 0.606689 |
| Transport injuries | 90-94 | 0.003200 | 0.000163 | 0.629363 | 0.005043 | 0.000388 | 0.506270 |
| Transport injuries | 95+ | 0.003049 | 0.000173 | 0.624384 | 0.006148 | 0.000440 | 0.401822 |
| Unintentional injuries | 0-4 | 0.293610 | 0.000037 | 0.006841 | 0.280398 | 0.000043 | 0.007234 |
| Unintentional injuries | 5-9 | 0.313929 | 0.000015 | 0.012083 | 0.419744 | 0.000020 | 0.009950 |
| Unintentional injuries | 10-14 | 0.292908 | 0.000004 | 0.019419 | 0.482889 | 0.000015 | 0.014427 |
| Unintentional injuries | 15-19 | 0.222639 | 0.000007 | 0.029082 | 0.462979 | 0.000030 | 0.020499 |
| Unintentional injuries | 20-24 | 0.191426 | 0.000009 | 0.038728 | 0.419989 | 0.000046 | 0.028722 |
| Unintentional injuries | 25-29 | 0.177634 | 0.000010 | 0.049257 | 0.363228 | 0.000053 | 0.040278 |

| | | | | | | | |
|---|---|---|---|---|---|---|---|
| Unintentional injuries | 30-34 | 0.163694 | 0.000011 | 0.062442 | 0.302600 | 0.000054 | 0.055382 |
| Unintentional injuries | 35-39 | 0.144086 | 0.000013 | 0.080450 | 0.249828 | 0.000053 | 0.074340 |
| Unintentional injuries | 40-44 | 0.123369 | 0.000016 | 0.102931 | 0.208732 | 0.000057 | 0.097192 |
| Unintentional injuries | 45-49 | 0.106951 | 0.000020 | 0.127124 | 0.177223 | 0.000065 | 0.122749 |
| Unintentional injuries | 50-54 | 0.097349 | 0.000026 | 0.149700 | 0.155949 | 0.000079 | 0.148157 |
| Unintentional injuries | 55-59 | 0.093551 | 0.000033 | 0.167943 | 0.143211 | 0.000095 | 0.171605 |
| Unintentional injuries | 60-64 | 0.095225 | 0.000042 | 0.177657 | 0.138071 | 0.000117 | 0.188233 |
| Unintentional injuries | 65-69 | 0.107937 | 0.000064 | 0.174267 | 0.143510 | 0.000152 | 0.192524 |
| Unintentional injuries | 70-74 | 0.147111 | 0.000153 | 0.157940 | 0.168328 | 0.000258 | 0.182614 |
| Unintentional injuries | 75-79 | 0.223998 | 0.000452 | 0.137930 | 0.223952 | 0.000642 | 0.162985 |
| Unintentional injuries | 80-84 | 0.339298 | 0.001216 | 0.125251 | 0.312783 | 0.001590 | 0.144979 |
| Unintentional injuries | 85-89 | 0.488592 | 0.003179 | 0.120320 | 0.424667 | 0.003852 | 0.135173 |
| Unintentional injuries | 90-94 | 0.671524 | 0.006402 | 0.119892 | 0.557304 | 0.007496 | 0.131049 |
| Unintentional injuries | 95+ | 0.887921 | 0.010881 | 0.123982 | 0.710520 | 0.012511 | 0.132820 |

For acute alcohol-related causes, incidence, (years lived with disability (YLD) and mortality rates are from the Global Burden of Disease data.[3]

#### Costs for alcohol-related diseases and injuries

*Table S4: Annual cost per prevalent or incident case of alcohol-related diseases and injuries*

| Disease | Unit costed | Sex | Unit Cost in AUD | | |
|---|---|---|---|---|---|
| | | | 0 to 34 years | 35 to 64 years | 65+ years |
| Alcoholic cardiomyopathy | Prevalent case | female | 6,237 | 4,540 | 2,190 |
| Alcoholic cardiomyopathy | Prevalent case | male | 6,800 | 4,849 | 2,703 |
| Alcohol use disorders | Incident case | female | 1,267 | 3,135 | 2,634 |
| Alcohol use disorders | Incident case | male | 1,028 | 2,843 | 3,306 |
| Atrial fibrillation and flutter | Prevalent case | female | 58,429 | 2,019 | 2,148 |
| Atrial fibrillation and flutter | Prevalent case | male | 50,753 | 2,480 | 2,645 |
| Breast cancer | Incident case | female | 89,110 | 96,154 | 64,665 |
| Cirrhosis | Prevalent case | female | 31 | 65 | 62 |
| Cirrhosis | Prevalent case | male | 19 | 56 | 61 |
| Colon and rectum cancer | Incident case | female | 67,871 | 57,662 | 50,905 |
| Colon and rectum cancer | Incident case | male | 66,273 | 62,028 | 52,405 |
| Diabetes mellitus type 2 | Prevalent case | female | 1,193 | 1,052 | 1,111 |
| Diabetes mellitus type 2 | Prevalent case | male | 972 | 1,280 | 1,459 |
| Epilepsy | Prevalent case | female | 3,675 | 4,414 | 2,655 |
| Epilepsy | Prevalent case | male | 3,901 | 5,892 | 3,540 |
| Hypertensive heart disease | Prevalent case | female | 10,206 | 5,113 | 1,132 |
| Hypertensive heart disease | Prevalent case | male | 4,758 | 2,600 | 1,420 |
| Interpersonal violence | Incident case | female | 1,956 | 5,893 | 2,720 |
| Interpersonal violence | Incident case | male | 1,141 | 3,135 | 2,914 |
| Intracerebral haemorrhage | Prevalent case | female | 1,216 | 1,955 | 2,996 |
| Intracerebral haemorrhage | Prevalent case | male | 2,393 | 3,223 | 5,099 |
| Ischaemic heart disease | Prevalent case | female | 1,417 | 2,434 | 1,969 |
| Ischaemic heart disease | Prevalent case | male | 1,041 | 2,123 | 1,832 |
| Ischaemic stroke | Prevalent case | female | 1,216 | 1,955 | 2,996 |
| Ischaemic stroke | Prevalent case | male | 2,393 | 3,223 | 5,099 |
| Larynx and pharynx cancer | Incident case | female | 46,692 | 43,210 | 30,063 |
| Larynx and pharynx cancer | Incident case | male | 61,199 | 42,924 | 37,603 |
| Lip and oral cavity cancer | Incident case | female | 33,861 | 80,096 | 56,344 |
| Lip and oral cavity cancer | Incident case | male | 39,837 | 55,421 | 47,585 |
| Liver cancer | Incident case | female | 104,565 | 70,109 | 43,928 |
| Liver cancer | Incident case | male | 127,378 | 67,322 | 68,287 |
| Lower respiratory infections | Incident case | female | 3,270 | 2,726 | 2,475 |
| Lower respiratory infections | Incident case | male | 3,177 | 2,186 | 2,439 |
| Oesophageal cancer | Incident case | female | 29,678 | 76,018 | 49,633 |
| Oesophageal cancer | Incident case | male | 16,251 | 49,465 | 50,102 |
| Pancreatitis | Prevalent case | female | 24,963 | 20,744 | 14,792 |
| Pancreatitis | Prevalent case | male | 36,366 | 28,155 | 17,509 |
| Self-harm | Incident case | female | 10,567 | 10,081 | 17,609 |
| Self-harm | Incident case | male | 7,785 | 11,069 | 21,763 |
| Transport injuries | Incident case | female | 5,023 | 8,202 | 11,914 |
| Transport injuries | Incident case | male | 7,757 | 10,796 | 15,166 |
| Unintentional injuries | Incident case | female | 684 | 2,197 | 3,771 |
| Unintentional injuries | Incident case | male | 648 | 1,638 | 3,645 |

Cost data from Australian Institute of Health and Welfare health expenditure data.[6,7] An average of the AIHW 2018/19 and the 2019/20 reported costs was used. The cost per case reported in this table is for the 'all areas' category in the AIHW report. The 'all areas' category reflects total costs for on health care costs for private hospital services, public hospital admitted patients, public hospital emergency department, public hospital outpatients, general practitioner services, specialist services, medical imaging, pathology, allied health and other services, and the pharmaceutical benefits scheme.

*Table S5: Average annual cost of all other diseases in the population**

| 5-year age groups | Cost per person in AUD | |
|---|---|---|
| | Female | Male |
| Under 5 years | 3,241 | 3,867 |
| 5-9 years | 1,201 | 1,480 |
| 10-14 years | 1,384 | 1,471 |
| 15-19 years | 2,510 | 1,839 |
| 20-24 years | 3,267 | 1,962 |
| 25-29 years | 4,232 | 2,061 |
| 30-34 years | 5,240 | 2,294 |
| 35-39 years | 4,866 | 2,707 |
| 40-44 years | 4,321 | 3,192 |
| 45-49 years | 4,417 | 3,854 |
| 50-54 years | 4,973 | 4,770 |
| 55-59 years | 5,684 | 5,984 |
| 60-64 years | 6,792 | 7,829 |
| 65-69 years | 8,414 | 10,043 |
| 70-74 years | 10,498 | 12,764 |
| 75-79 years | 12,912 | 15,890 |
| 80-84 years | 15,020 | 18,553 |
| 85 years and over | 16,556 | 20,896 |

Cost data from Australian Institute of Health and Welfare health expenditure data.[6,7]
* These are costs calculated for all other diseases not modelled. The costs are incurred in added years of life since a reduction of alcohol consumption prolongs life (See section in main manuscript titled *Development of the model structure and input data sources)*

#### Relative risks

*Table S6: Relative risk measures of disease and injury due to alcohol intake*

| Cause | Sex | g/day* | Mean | SD | Cause | Sex | g/day | Mean | SD |
|---|---|---|---|---|---|---|---|---|---|
| Atrial fibrillation and flutter | both | 72 | 1.535 | 0.0969 | Ischemic stroke | male | 72 | 1.451 | 0.1179 |
| Atrial fibrillation and flutter | both | 60 | 1.411 | 0.0788 | Ischemic stroke | male | 60 | 1.312 | 0.0776 |
| Atrial fibrillation and flutter | both | 48 | 1.312 | 0.0482 | Ischemic stroke | male | 48 | 1.159 | 0.0952 |
| Atrial fibrillation and flutter | both | 36 | 1.214 | 0.0370 | Ischemic stroke | male | 36 | 1.057 | 0.0666 |
| Atrial fibrillation and flutter | both | 24 | 1.131 | 0.0349 | Ischemic stroke | male | 24 | 0.97 | 0.0577 |
| Atrial fibrillation and flutter | both | 12 | 1.066 | 0.0173 | Ischemic stroke | male | 12 | 0.938 | 0.0571 |
| Atrial fibrillation and flutter | both | 0 | 1 | 0 | Ischemic stroke | male | 0 | 1 | 0 |
| Breast cancer | both | 72 | 1.476 | 0.1043 | Ischemic stroke | female | 72 | 1.43 | 0.1592 |
| Breast cancer | both | 60 | 1.452 | 0.0732 | Ischemic stroke | female | 60 | 1.3 | 0.0957 |
| Breast cancer | both | 48 | 1.443 | 0.0495 | Ischemic stroke | female | 48 | 1.145 | 0.1054 |
| Breast cancer | both | 36 | 1.433 | 0.0612 | Ischemic stroke | female | 36 | 0.985 | 0.0804 |
| Breast cancer | both | 24 | 1.329 | 0.0464 | Ischemic stroke | female | 24 | 0.85 | 0.0661 |
| Breast cancer | both | 12 | 1.17 | 0.0469 | Ischemic stroke | female | 12 | 0.824 | 0.0564 |
| Breast cancer | both | 0 | 1 | 0 | Ischemic stroke | female | 0 | 1 | 0 |
| Cirrhosis and other chronic liver diseases due to alcohol use | both | 72 | 9.427 | 1.9574 | Larynx cancer | both | 72 | 2.461 | 0.3750 |
| Cirrhosis and other chronic liver diseases due to alcohol use | both | 60 | 6.274 | 1.3676 | Larynx cancer | both | 60 | 2.144 | 0.3763 |
| Cirrhosis and other chronic liver diseases due to alcohol use | both | 48 | 4.673 | 0.8844 | Larynx cancer | both | 48 | 1.813 | 0.2860 |
| Cirrhosis and other chronic liver diseases due to alcohol use | both | 36 | 3.274 | 0.5551 | Larynx cancer | both | 36 | 1.531 | 0.2385 |
| Cirrhosis and other chronic liver diseases due to alcohol use | both | 24 | 2.055 | 0.2977 | Larynx cancer | both | 24 | 1.304 | 0.1666 |
| Cirrhosis and other chronic liver diseases due to alcohol use | both | 12 | 1.243 | 0.1704 | Larynx cancer | both | 12 | 1.12 | 0.1232 |
| Cirrhosis and other chronic liver diseases due to alcohol use | both | 0 | 1 | 0 | Larynx cancer | both | 0 | 1 | 0 |
| Colon and rectum cancer | both | 72 | 1.616 | 0.1227 | Lip and oral cavity cancer | both | 72 | 4.858 | 0.5959 |
| Colon and rectum cancer | both | 60 | 1.468 | 0.0730 | Lip and oral cavity cancer | both | 60 | 3.766 | 0.5258 |
| Colon and rectum cancer | both | 48 | 1.323 | 0.0880 | Lip and oral cavity cancer | both | 48 | 2.991 | 0.4115 |

| | | | | |
|---|---|---|---|---|
| Colon and rectum cancer | both | 36 | 1.237 | 0.0480 |
| Colon and rectum cancer | both | 24 | 1.156 | 0.0462 |
| Colon and rectum cancer | both | 12 | 1.078 | 0.0230 |
| Colon and rectum cancer | both | 0 | 1 | 0 |
| Diabetes mellitus | male | 72 | 1.198 | 0.0694 |
| Diabetes mellitus | male | 60 | 1.165 | 0.0878 |
| Diabetes mellitus | male | 48 | 1.084 | 0.0781 |
| Diabetes mellitus | male | 36 | 1 | 0.0582 |
| Diabetes mellitus | male | 24 | 0.932 | 0.0482 |
| Diabetes mellitus | male | 12 | 0.921 | 0.0464 |
| Diabetes mellitus | male | 0 | 1 | 0 |
| Diabetes mellitus | female | 72 | 1.172 | 0.2148 |
| Diabetes mellitus | female | 60 | 1.074 | 0.1730 |
| Diabetes mellitus | female | 48 | 0.945 | 0.1112 |
| Diabetes mellitus | female | 36 | 0.836 | 0.0712 |
| Diabetes mellitus | female | 24 | 0.76 | 0.0541 |
| Diabetes mellitus | female | 12 | 0.733 | 0.0429 |
| Diabetes mellitus | female | 0 | 1 | 0 |
| Epilepsy | both | 72 | 2.480 | 0.3099 |
| Epilepsy | both | 60 | 2.186 | 0.2145 |
| Epilepsy | both | 48 | 1.872 | 0.2375 |
| Epilepsy | both | 36 | 1.585 | 0.1518 |
| Epilepsy | both | 24 | 1.353 | 0.1314 |
| Epilepsy | both | 12 | 1.177 | 0.0656 |
| Epilepsy | both | 0 | 1 | 0 |
| Hypertensive heart disease | both | 72 | 1.860 | 0.2329 |
| Hypertensive heart disease | both | 60 | 1.705 | 0.2240 |
| Hypertensive heart disease | both | 48 | 1.614 | 0.2038 |
| Hypertensive heart disease | both | 36 | 1.479 | 0.1344 |
| Hypertensive heart disease | both | 24 | 1.315 | 0.0995 |
| Hypertensive heart disease | both | 12 | 1.046 | 0.0727 |
| Hypertensive heart disease | both | 0 | 1 | 0 |
| Interpersonal violence | both | 72 | 1.516 | 0.1561 |
| Interpersonal violence | both | 60 | 1.452 | 0.1286 |
| Interpersonal violence | both | 48 | 1.396 | 0.1584 |
| Interpersonal violence | both | 36 | 1.345 | 0.1135 |
| Interpersonal violence | both | 24 | 1.256 | 0.1033 |

| | | | | |
|---|---|---|---|---|
| Lip and oral cavity cancer | both | 36 | 2.311 | 0.2990 |
| Lip and oral cavity cancer | both | 24 | 1.738 | 0.1985 |
| Lip and oral cavity cancer | both | 12 | 1.293 | 0.1212 |
| Lip and oral cavity cancer | both | 0 | 1 | 0 |
| Liver cancer due to alcohol use | both | 72 | 1.424 | 0.1957 |
| Liver cancer due to alcohol use | both | 60 | 1.372 | 0.1528 |
| Liver cancer due to alcohol use | both | 48 | 1.31 | 0.1538 |
| Liver cancer due to alcohol use | both | 36 | 1.225 | 0.1138 |
| Liver cancer due to alcohol use | both | 24 | 1.14 | 0.1084 |
| Liver cancer due to alcohol use | both | 12 | 1.067 | 0.0691 |
| Liver cancer due to alcohol use | both | 0 | 1 | 0 |
| Lower respiratory infections | both | 72 | 1.357 | 0.1365 |
| Lower respiratory infections | both | 60 | 1.226 | 0.0987 |
| Lower respiratory infections | both | 48 | 1.127 | 0.0997 |
| Lower respiratory infections | both | 36 | 1.064 | 0.0742 |
| Lower respiratory infections | both | 24 | 1.026 | 0.0679 |
| Lower respiratory infections | both | 12 | 1.013 | 0.0339 |
| Lower respiratory infections | both | 0 | 1 | 0 |
| Nasopharynx cancer | both | 72 | 4.545 | 0.2250 |
| Nasopharynx cancer | both | 60 | 3.803 | 0.1513 |
| Nasopharynx cancer | both | 48 | 3.062 | 0.0982 |
| Nasopharynx cancer | both | 36 | 2.385 | 0.0770 |
| Nasopharynx cancer | both | 24 | 1.839 | 0.0349 |
| Nasopharynx cancer | both | 12 | 1.371 | 0.0145 |
| Nasopharynx cancer | both | 0 | 1 | 0 |
| Oesophageal cancer | both | 72 | 2.669 | 0.3250 |
| Oesophageal cancer | both | 60 | 2.452 | 0.3033 |
| Oesophageal cancer | both | 48 | 2.202 | 0.2482 |
| Oesophageal cancer | both | 36 | 1.815 | 0.1923 |
| Oesophageal cancer | both | 24 | 1.466 | 0.1416 |
| Oesophageal cancer | both | 12 | 1.212 | 0.1041 |
| Oesophageal cancer | both | 0 | 1 | 0 |
| Other pharynx cancer | both | 72 | 4.764 | 0.8319 |
| Other pharynx cancer | both | 60 | 3.972 | 0.6482 |
| Other pharynx cancer | both | 48 | 3.199 | 0.5625 |
| Other pharynx cancer | both | 36 | 2.519 | 0.3714 |
| Other pharynx cancer | both | 24 | 1.943 | 0.2594 |

| | | | | |
|---|---|---|---|---|
| Interpersonal violence | both | 12 | 1.129 | 0.0903 |
| Interpersonal violence | both | 0 | 1 | 0 |
| Intracerebral haemorrhage | male | 72 | 1.971 | 0.1666 |
| Intracerebral haemorrhage | male | 60 | 1.705 | 0.1380 |
| Intracerebral haemorrhage | male | 48 | 1.458 | 0.1495 |
| Intracerebral haemorrhage | male | 36 | 1.31 | 0.1107 |
| Intracerebral haemorrhage | male | 24 | 1.162 | 0.1051 |
| Intracerebral haemorrhage | male | 12 | 1.068 | 0.0686 |
| Intracerebral haemorrhage | male | 0 | 1 | 0 |
| Intracerebral haemorrhage | female | 72 | 2.276 | 0.3145 |
| Intracerebral haemorrhage | female | 60 | 1.964 | 0.2367 |
| Intracerebral haemorrhage | female | 48 | 1.614 | 0.2048 |
| Intracerebral haemorrhage | female | 36 | 1.337 | 0.1528 |
| Intracerebral haemorrhage | female | 24 | 1.11 | 0.1232 |
| Intracerebral haemorrhage | female | 12 | 1.031 | 0.0722 |
| Intracerebral haemorrhage | female | 0 | 1 | 0 |
| Ischemic heart disease | male | 72 | 1.091 | 0.0862 |
| Ischemic heart disease | male | 60 | 0.993 | 0.0566 |
| Ischemic heart disease | male | 48 | 0.906 | 0.0607 |
| Ischemic heart disease | male | 36 | 0.871 | 0.0449 |
| Ischemic heart disease | male | 24 | 0.857 | 0.0418 |
| Ischemic heart disease | male | 12 | 0.865 | 0.0403 |
| Ischemic heart disease | male | 0 | 1 | 0 |
| Ischemic heart disease | female | 72 | 1.107 | 0.1140 |
| Ischemic heart disease | female | 60 | 1.012 | 0.0778 |
| Ischemic heart disease | female | 48 | 0.932 | 0.0834 |
| Ischemic heart disease | female | 36 | 0.882 | 0.0551 |
| Ischemic heart disease | female | 24 | 0.846 | 0.0508 |
| Ischemic heart disease | female | 12 | 0.823 | 0.0492 |
| Ischemic heart disease | female | 0 | 1 | 0 |
| Other pharynx cancer | both | 12 | 1.472 | 0.1296 |
| Other pharynx cancer | both | 0 | 1 | 0 |
| Pancreatitis | both | 72 | 3.298 | 0.5064 |
| Pancreatitis | both | 60 | 2.217 | 0.5036 |
| Pancreatitis | both | 48 | 1.717 | 0.3260 |
| Pancreatitis | both | 36 | 1.471 | 0.2446 |
| Pancreatitis | both | 24 | 1.228 | 0.2031 |
| Pancreatitis | both | 12 | 1.073 | 0.1760 |
| Pancreatitis | both | 0 | 1 | 0 |
| Self-harm | both | 72 | 1.927 | 0.3232 |
| Self-harm | both | 60 | 1.734 | 0.2597 |
| Self-harm | both | 48 | 1.545 | 0.2337 |
| Self-harm | both | 36 | 1.376 | 0.1788 |
| Self-harm | both | 24 | 1.23 | 0.1431 |
| Self-harm | both | 12 | 1.107 | 0.1110 |
| Self-harm | both | 0 | 1 | 0 |
| Transport injuries | both | 72 | 1.552 | 0.2120 |
| Transport injuries | both | 60 | 1.456 | 0.1612 |
| Transport injuries | both | 48 | 1.366 | 0.1508 |
| Transport injuries | both | 36 | 1.288 | 0.1135 |
| Transport injuries | both | 24 | 1.22 | 0.0862 |
| Transport injuries | both | 12 | 1.163 | 0.0829 |
| Transport injuries | both | 0 | 1 | 0 |
| Unintentional injuries | both | 72 | 1.266 | 0.1255 |
| Unintentional injuries | both | 60 | 1.221 | 0.1023 |
| Unintentional injuries | both | 48 | 1.182 | 0.1031 |
| Unintentional injuries | both | 36 | 1.168 | 0.0747 |
| Unintentional injuries | both | 24 | 1.154 | 0.0696 |
| Unintentional injuries | both | 12 | 1.09 | 0.0436 |
| Unintentional injuries | both | 0 | 1 | 0 |

Relative risk measures do not include alcohol use disorders, which is entirely attributable to alcohol use, so we adjust the rates in proportion to the change in alcohol. In the absence of data, we assume the 'curve' flattens off after the indicated g/day limit e.g. for Atrial fibrillation and flutter, above 72g/d, we use the RR for 72 g/d. g/day: grams per day, SD: standard deviation

#### A schematic description of The Alcohol Policy (TAP) Model

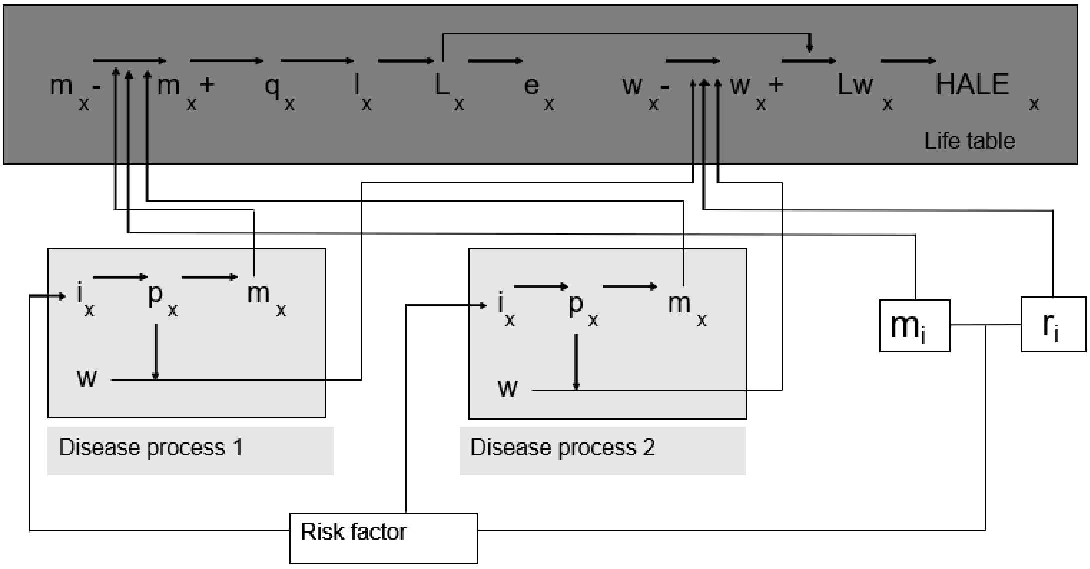

*Figure S2: Schematic description of the proportional multistate lifetable including the interaction with the disease models and the risk factor.*[8]

All of the factors are age-specific, with the x in subscript denoting age. The areas highlighted in yellow are the disease models while the grey is the lifetable. i is incidence, p is prevalence, m is mortality rate, w is disability weight (proportion of quality of life considered to be lost due to disease or disability), q is mortality probability, l is number of survivors, L is life years, e is life expectancy, Lwx is number of disability adjusted life years lived in age interval x, HALE = health-adjusted life expectancy. '-'denotes parameter related to diseases or causes that specifically excludes modelled diseases or injuries and '+' relates to all modelled diseases in the model.  A change in the risk factor exposure (alcohol use) translates into changes in incidence (ix), which changes disease-specific prevalence (px) and mortality (mx). For presentation purposes, only two diseases processes are shown. A change in the risk factor exposure (alcohol use) impacts modelled injuries: fatalities (mi) impact directly on mortality and non-fatal injuries (ri) impact on years lived with disability.

#### Additional results from main analysis

##### Changes in incident cases of disease and injuries

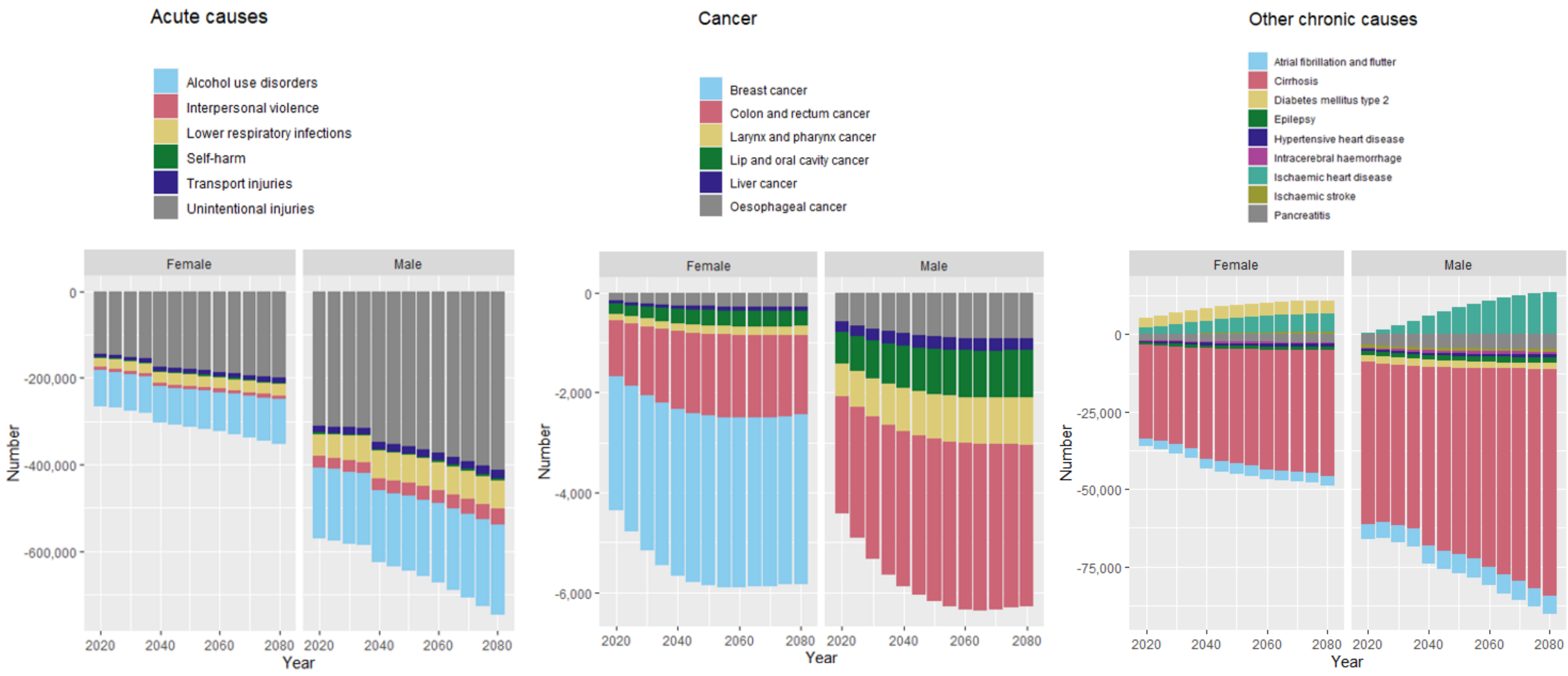

*Figure S3: Changes in incident cases if alcohol consumption was eliminated in the Australian population (main analysis)*

Negative values can be explained by the questionable alcohol protective effect in the relative risk measures and second, disease & injury cases that arise in added years of life (people live longer due to reduced alcohol harm and they are still at risk of these diseases and injuries).

#### Changes in mortality

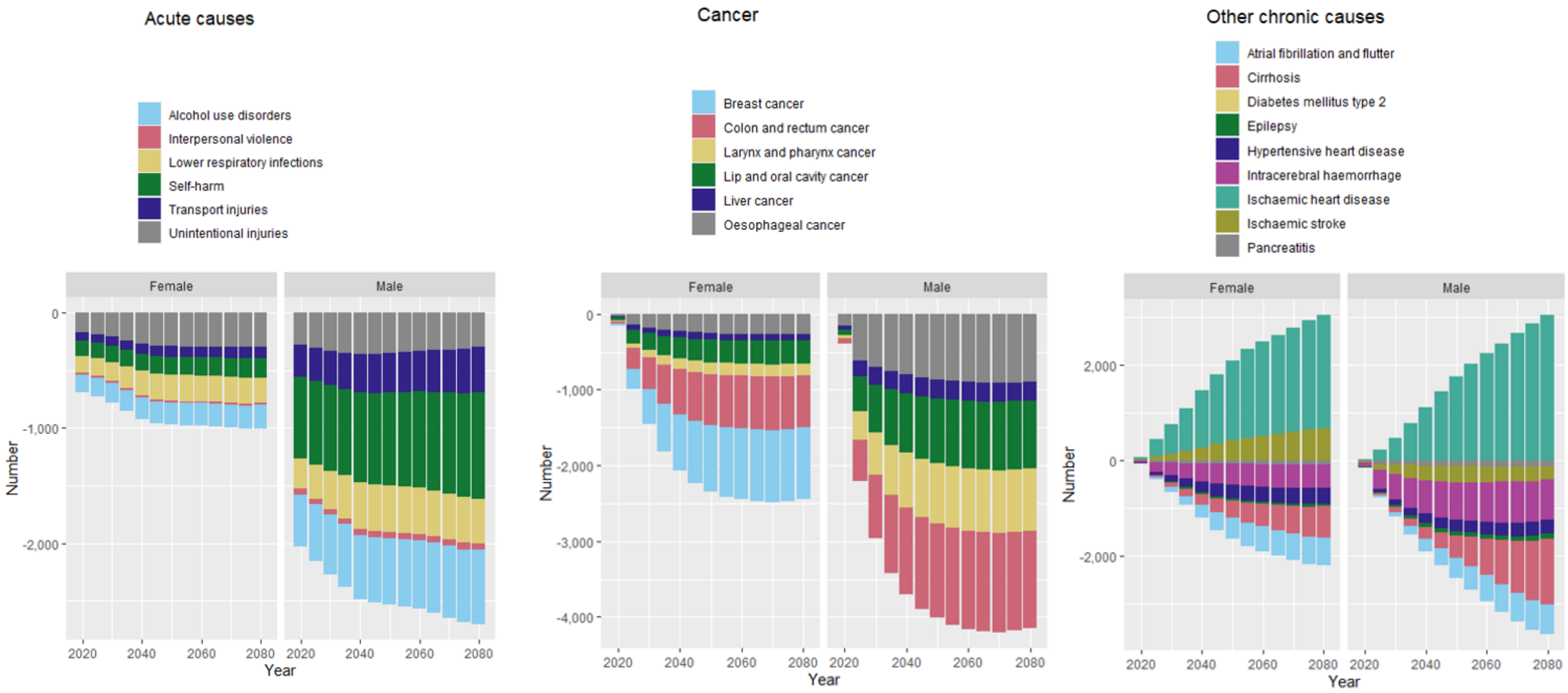

*Figure S4: Changes in mortality over time if alcohol consumption was eliminated in the Australian population (main analysis)*

Negative values can be explained by the questionable alcohol protective effect and cases that arise when people live longer due to reduced alcohol harm and they are still at risk of mortality from these diseases and injuries. The majority of diabetes-related deaths are from ischemic heart disease and ischemic stroke. Therefore, we ignore diabetes mellitus type 2 mortality in the model, assuming it was covered by ischaemic heart disease and ischaemic stroke.

### Additional results from sensitivity analysis

#### Changes in incidence and mortality

*Table S7: Changes in incidence and mortality over the first 25 years when the protective effects of low and moderate alcohol consumption are excluded*

| **Incident cases that could be prevented** | | | |
|---|---|---|---|
| | **Female (Mean, 95%UIs)** | **Male (Mean, 95%UIs)** | **Total (Mean, 95%UIs)** |
| **Sensitivity analysis (with protective effects of low, moderate alcohol consumption on ischemic heart disease, diabetes mellitus type 2 and Ischemic stroke excluded)[a]** | | | |
| Diabetes mellitus type 2 | 29,764 | 86,190 | 115,954 |
| | (-2,946 to 111,513) | (-7,899 to 220,580) | (-1,614 to 268,541) |
| Ischaemic heart disease | 23,150 | 161,784 | 184,934 |
| | (-14,390 to 89,480) | (-69,543 to 478,130) | (-51,351 to 498,010) |
| Ischaemic stroke | 16,308 | 29,097 | 45,405 |
| | (-1,482 to 39,972) | (5,800 to 57,472) | (15,673 to 82,159) |
| **Main analysis (with protective effects of low, moderate alcohol consumption on ischemic heart disease, diabetes mellitus type 2 and Ischemic stroke included)[b]** | | | |
| Diabetes mellitus type 2 | -96,470 | 59,579 | -36,891 |
| | (-266,530 to 78,705) | (-115,782 to 225,786) | (-279,513 to 208,483) |
| Ischaemic heart disease | -86,736 | -99,370 | -186,106 |
| | (-242,660 to 70,846) | (-650,104 to 425,468) | (-744,135 to 350,779) |
| Ischaemic stroke | -7,816 | 24,497 | 16,680 |
| | (-56,953 to 39,445) | (-12,518 to 59,309) | (-44,829 to 74,104) |
| **Deaths that could be averted** | | | |
| | **Female (Mean, 95%UIs)** | **Male (Mean, 95%UIs)** | **Total (Mean, 95%UIs)** |
| **Sensitivity analysis (with protective effects of low, moderate alcohol consumption on ischemic heart disease, diabetes mellitus type 2 and Ischemic stroke excluded)[a]** | | | |
| Diabetes mellitus type 2 | 0 | 0 | 0 |
| | (0 to 0) | (0 to 0) | (0 to 0) |
| Ischaemic heart disease | -705 | -482 | -1,186 |
| | (-7,513 to 11,291) | (-15,012 to 20,502) | (-18,544 to 21,498) |
| Ischaemic stroke | 3,187 | 7,190 | 10,378 |
| | (-2,629 to 11,070) | (-821 to 17,276) | (410 to 23,144) |
| **Main analysis (with protective effects of low, moderate alcohol consumption on ischemic heart disease, diabetes mellitus type 2 and Ischemic stroke included)[b]** | | | |
| Diabetes mellitus type 2[#] | 0 | 0 | 0 |
| | (0 to 0) | (0 to 0) | (0 to 0) |
| Ischaemic heart disease | -19,731 | -16,888 | -36,619 |
| | (-47,157 to 8,808) | (-50,986 to 16,649) | (-80,506 to 5,453) |
| Ischaemic stroke | -4,308 | 5,841 | 1,534 |
| | (-19,955 to 11,248) | (-7,024 to 18,258) | (-19,130 to 21,171) |

[a]Sensitivity analysis: Alcohol consumption eliminated in the Australian population with protective effects of low, moderate alcohol consumption on ischemic heart disease, diabetes mellitus type 2 and Ischemic stroke excluded.
[b]Main analysis: Alcohol consumption eliminated in the Australian population with protective effects of low, moderate alcohol consumption on ischemic heart disease, diabetes mellitus type 2 and Ischemic stroke included.
[#]The majority of diabetes-related deaths are from ischemic heart disease and ischemic stroke. Therefore, we ignore diabetes mellitus type 2 mortality in the model, assuming it was covered by ischaemic heart disease and ischaemic stroke.
Negative values can be explained by the questionable alcohol protective effect and cases that arise when people live longer due to reduced alcohol harm

#### Changes in Health adjusted life years (HALYs)

*Table S8: Sex specific HALYs gained for various scenarios modelled*

| Scenario | Age group (years) | HALYs in Millions, mean (95%UIs) | | |
|---|---|---|---|---|
| | | Female | Male | Total |
| Avoidable burden (base case) | 0-25 | 1.5 (1 to 2.1) | 3.5 (2.5 to 4.5) | 5.1 (4 to 6.2) |
| | 0-60 | 5 (3.2 to 6.7) | 12 (9.1 to 15) | 17 (14 to 21) |
| Avoidable burden with protective effects excluded | 0-25 | 1.8 (1.4 to 2.3) | 3.7 (2.8 to 4.6) | 5.5 (4.6 to 6.5) |
| | 0-60 | 6.3 (5 to 7.7) | 13 (10 to 15) | 19 (16 to 22) |
| Met alcohol consumption guidelines | 0-25 | 1.2 (0.7 to 1.8) | 3.2 (2.2 to 4.1) | 4.4 (3.2 to 5.5) |
| | 0-60 | 4.2 (2.4 to 5.9) | 11 (8.1 to 14) | 15 (12 to 19) |

Scenarios modelled: Avoidable burden: Population level elimination of alcohol consumption (Base case); Avoidable burden with protective effects excluded: Population level elimination of alcohol consumption with protective effects excluded (assessing sensitivity to excluding the beneficial/protective effects of low and moderate alcohol consumption on ischemic heart disease, diabetes mellitus type 2 and Ischemic stroke)[9]; Met guidelines: Population met the current alcohol consumption guidelines (no more than 10 standard drinks a week).[10]

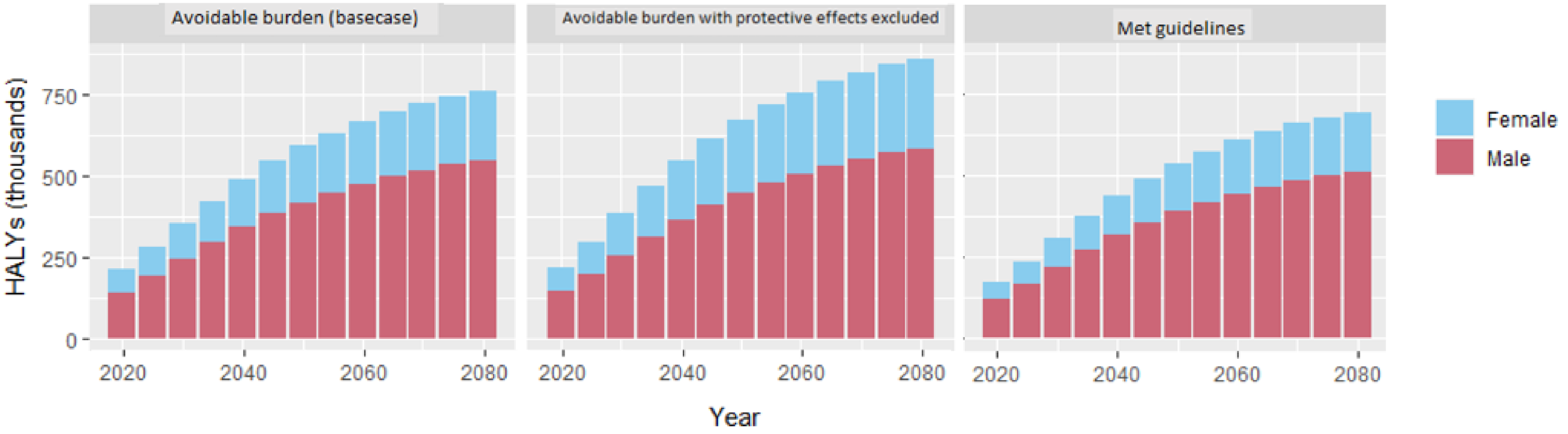

*Figure S5: HALYs gained over different time periods for various modelled scenarios*

Scenarios modelled: Avoidable burden: Population level elimination of alcohol consumption (Base case); Avoidable burden with protective effects excluded: Population level elimination of alcohol consumption with protective effects excluded (assessing sensitivity to excluding the beneficial/protective effects of low and moderate alcohol consumption on ischemic heart disease, diabetes mellitus type 2 and Ischemic stroke)[9]; Met guidelines: Population met the current alcohol consumption guidelines (no more than 10 standard drinks a week).[10]

#### Changes in healthcare costs by area of expenditure

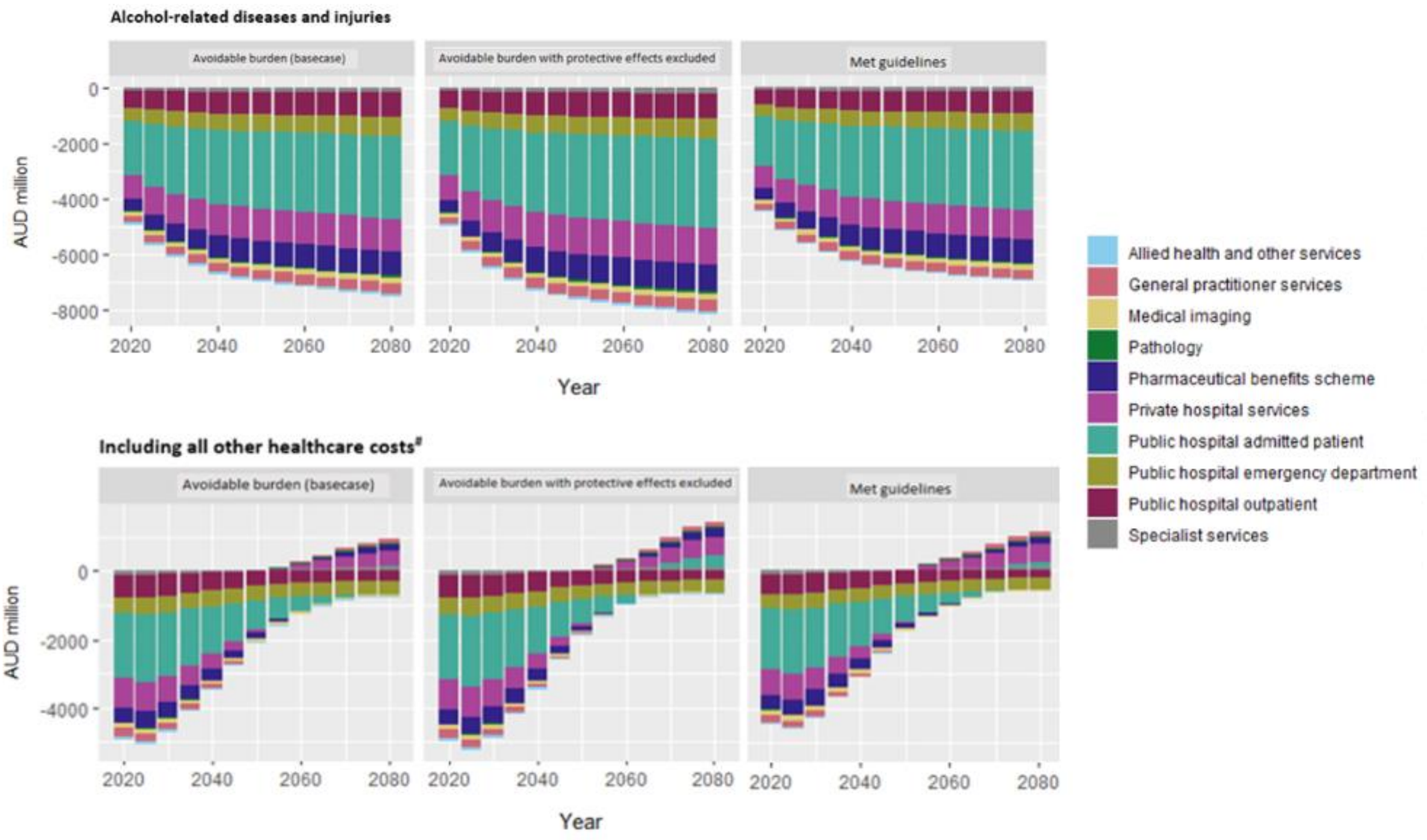

*Figure S6: changes in healthcare costs over different time periods for four modelled scenarios*

The estimated changes are presented per the areas of expenditure defined by the Australian Institute of Health and Welfare.[6,7] Chart titled *alcohol related diseases and injuries* shows changes for when only the changes in the costs of alcohol-related diseases and injuries are modelled. #Chart titled i*ncluding all other healthcare costs* shows changes for both the costs of alcohol-related diseases and injuries and the costs of all other health care due to changes in life expectancy, (i.e., costs incurred in added years of life since a reduction of alcohol consumption prolongs life).[11,12] Scenarios modelled: Avoidable burden: Population level elimination of alcohol consumption (Base case/main analysis); Avoidable burden with protective effects excluded: Population level elimination of alcohol consumption with protective effects excluded (assessing sensitivity to excluding the beneficial/protective effects of low and moderate alcohol consumption on ischemic heart disease, diabetes mellitus type 2 and Ischemic stroke)[9]; Met guidelines: Population met the current alcohol consumption guidelines (no more than 10 standard drinks a week).[10]
